# Causal effects of circulating cytokine concentrations on risk of Alzheimer’s disease: A bidirectional two-sample Mendelian randomization study

**DOI:** 10.1101/2020.11.18.20232629

**Authors:** Panagiota Pagoni, Laura D Howe, George Davey Smith, Yoav Ben-Shlomo, Evie Stergiakouli, Emma L Anderson

**Author notes:** Correspondence to Panagiota Pagoni, MRC Integrative Epidemiology Unit, Oakfield House, Oakfield Grove, Bristol, United Kingdom, BS8 2BN. joint last authors (Evie Stergiakouli, Emma L Anderson). **Competing interests:** None declared.

## Abstract

**Background:** There is considerable interest in the role of neuroinflammation in the pathogenesis of Alzheimer’s disease. Evidence from observational studies suggests an association between cytokine concentrations and Alzheimer’s disease. However, establishing a causal role of cytokine concentrations on risk of Alzheimer’s disease is challenging due to bias from reverse causation and residual confounding.

**Methods:** We used two-sample MR to explore causal effects of circulating cytokine concentrations on Alzheimer’s disease and vice versa, employing genetic variants associated with cytokine concentrations (N=8,293) and Alzheimer’s disease (71,880 cases / 383,378 controls) from the largest non-overlapping genome-wide association studies (GWAS) of European ancestry.

**Results:** There was weak evidence to suggest that 1 standard deviation (SD) increase in levels of CTACK (CCL27) (OR= 1.09 95%CI: 1.01 to 1.19, p=0.03) increased risk of Alzheimer’s disease. There was also weak evidence of a causal effect of 1 SD increase in levels of MIP-1b (CCL4) (OR=1.04 95%CI: 0.99 to 1.09, p=0.08), Eotaxin (OR=1.08 95%CI: 0.99 to 1.17, p =0.10), GROa (CXCL1) (OR=1.04 95%CI: 0.99 to 1.10, p=0.15), MIG (CXCL9) (OR=1.17 95%CI: 0.97 to 1.41, p=0.10), IL-8 (Wald Ratio: OR=1.21 95%CI: 0.97 to 1.51, p=0.09) and IL-2 (Wald Ratio: OR=1.21 95%CI: 0.94 to 1.56, p=0.14) on greater risk of Alzheimer’s disease. There was little evidence of a causal effect of genetic liability to Alzheimer’s disease on circulating cytokine concentrations.

**Conclusions:** Our study provides some evidence supporting a causal role of cytokines in the pathogenesis of Alzheimer’s disease. However, more studies are needed to elucidate the specific mechanistic pathways via which cytokines alter the risk of Alzheimer’s disease.

## INTRODUCTION

It has been estimated that 47 million people are affected by Alzheimer’s disease and other forms of dementia worldwide and this number is likely to increase, mostly due to ageing of the population (1). Thus, dementia is a major public health challenge and priority.

Over the last decade attention has been drawn to the interplay between the central nervous system (CNS) and immune responses (i.e. neuroinflammation) (2) in the pathogenesis of Alzheimer’s disease, mostly due to evidence stemming from observational studies suggesting that inflammatory disorders (e.g. rheumatoid arthritis) (3) and chronic inflammation (e.g. periodontitis) (4) are associated with a higher risk of Alzheimer’s disease. The neuroinflammation hypothesis suggests that in response to production and deposition of Aβ in the brain, the CNS activates microglia to protect the cells and overall brain function. As part of this defense mechanism, secondary inflammatory mediators such as cytokines (5, 6), lipid metabolites and free radicals are generated to rehabilitate the homeostasis and ensure a healthy neural function (7). However, over-activation of microglia may occur and lead to an exaggerated release of pro- and anti-inflammatory mediators (8), resulting in deterioration or even initiation of neurological diseases.

Many observational studies have examined the association between cytokine concentrations and risk of Alzheimer’s disease. A recent meta-analysis of 170 studies reported elevated peripheral levels of C-reactive protein (CRP), interleukin-6 (IL-6), interleukin-1 beta (IL-1β), soluble tumour necrosis factor receptor 1 and 2 (sTNFR-1 & sTNFR-2), interleukin-10 (IL-10), monocyte chemoattractant protein-1 (MCP-1) and transforming growth factor-1 (TGF-1) in individuals with an Alzheimer’s disease diagnosis compared to healthy controls (9). However, deciphering the role of inflammatory markers in the pathogenesis of Alzheimer’s disease is challenging because of the potential bias due to reverse causation, which refers to the possibility of Alzheimer’s disease being the cause rather than the consequence of inflammation. Confounding is another key source of bias inherent in existing observational studies, as Alzheimer’s disease patients tend to be older, and thus are more at risk of conditions such as obesity and hypertension, which may increase systemic inflammation.

A promising method that could help overcome the limitations of observational studies is Mendelian randomization (MR). Mendelian randomization uses genetic variants as proxies for an exposure and allows investigation of the causal effect of the exposure on the outcome of interest (10, 11). Moreover, two-sample MR studies, where the effect estimates of the genetic variants of exposure and outcome are extracted from different genome-wide association studies (GWAS) (12) can be useful, as they allow the estimation of causal effects without requiring the exposure and outcome to be measured in the same participants. Deciphering the causal role of neuroinflammation in the pathogenesis of Alzheimer’s disease could provide valuable information towards possible therapeutic targets. However, to date, very few studies have used MR to examine this question (13-16), and existing MR studies have included only a small, select group of previously implicated cytokines such as TNF-a, interleukins and CRP. Considering the plethora of observational evidence indicating a role of cytokines in the pathogenesis of Alzheimer’s disease and the limitations of observational studies to infer causality, we aimed to examine the bidirectional causal effects of cytokines on risk of Alzheimer’s disease in a MR framework.

## METHODS

### GWAS Summary Data

We used the most comprehensive and largest publicly available GWAS meta-analysis on concentrations of 41 circulating cytokines including up to 8,293 individuals from three independent population cohorts (The Cardiovascular Risk in Young Finns Study (YFS), FINRISK 1997 and FINRISK 2002)(17). Cytokines GWAS were adjusted for sex, age and body mass index (BMI).

For Alzheimer’s disease the most recently conducted GWAS was used, which consists of three phases. In phase one, 24,087 clinically diagnosed cases from the International Genomics of Alzheimer’s Project (IGAP), the Alzheimer’s Disease Sequencing Project (ADSP) and the Alzheimer’s disease working group of the Psychiatric Genomics Consortium (PGZ-ALZ) and 55,058 matched controls were included. In phase two, cases consist of 47,793 proxy-cases as defined in the UK Biobank and 328,320 proxy-controls (18). Participants were considered proxy-cases if they had positively responded to the question ‘Has your mother or father ever suffered from Alzheimer’s disease/Dementia’. Finally, phase three is a meta-analysis of all individuals in phase one & two and therefore consists of 71,880 cases and 383,378 controls of European ancestry. In our main analysis, we used data from phase one. As these summary data were not publicly available, we used summary estimates from Korologou-Linden et al. (19), which corresponds to phase one of the Alzheimer’s GWAS. Effect estimates were adjusted for sex, age, genotyping array and assessment centre. This study, assessed the effect of genetic liability to Alzheimer’s disease on circulating cytokine concentrations, as captured by common genetic variants. When examining binary exposures in MR settings, causal inferences are valid for the continuous liability underlying the binary exposure (20). Thus, when we test for the causal effect of Alzheimer’s disease, we are essentially examining the effect of genetic liability for this exposure which can be present in an individual even when they do not have a diagnosis.

### Instrument selection

For each exposure of interest, approximately independent genome-wide significant single nucleotide polymorphisms (SNPs) were identified (r^2^<0.01 within a 10,000 kb window, p<5×10^−08^). Eight cytokines had no SNPs available at the genome wide significance threshold, thus we relaxed the significance threshold to p<5×10^−07^ for these. When investigating the causal effects of cytokine concentrations on Alzheimer’s disease, we extracted the SD-scaled effect sizes and standard errors for cytokine SNPs from the publicly available cytokine GWAS, and corresponding log odds and standard errors from the Alzheimer’s disease GWAS. When investigating the reverse direction, we extracted log odds and standard errors for Alzheimer’s disease SNPs from the Alzheimer’s GWAS and corresponding SD-scaled effect sizes and standard errors from the cytokines GWAS.

Before estimating total causal effects, we harmonised the alleles of our datasets and further information about the procedure followed can be found in Supplementary Note 1.

### Mendelian randomization analysis

Univariable MR was employed to estimate the total causal effect of each circulating cytokine concentration on Alzheimer’s disease, and vice versa. MR relies on three assumptions that the genetic variants should satisfy to be considered as valid instruments and therefore yield unbiased causal effect estimates. Genetic variants i) must be strongly associated with the exposure of interest, ii) independently of any confounders of the exposure – outcome association and iii) they are associated with the outcome only via the exposure (i.e. no horizontal pleiotropy) (21).

When a single variant was available as a proxy for the exposure of interest, the Wald ratio estimator was employed to quantify the causal effect. When multiple variants were available, the Inverse-Variance-Weighted (IVW) method was used to estimate the total causal effect, which is equivalent to fitting a weighted linear regression of the gene-outcome associations on the gene-exposure associations, with the intercept term constrained to zero. Therefore, IVW estimates assume that all genetic variants are valid instruments with no pleiotropic effects (22, 23).

### Sensitivity analyses

We performed a series of sensitivity analyses to test the validity of the core assumptions which MR relies on (Table 1). A more comprehensive description of these sensitivity analyses can be found in Supplementary Note 2 and Supplementary Note 3.

**Table 1.** Description of sensitivity analyses conducted, and the theoretical context of each analysis.

| Sensitivity analysis | Theoretical context of each method | Reference |
| --- | --- | --- |
| F statistic | Genetic variant strongly associated with exposure of interest | (24) |
| MR-Egger | Adjusting for horizontal pleiotropic effects of genetic variants | (25) |
| Weighted median | 50% of the genetic instruments are invalid | (26) |
| Leave-one variant out | Pleiotropic effects of variants | (27) |
| Cohran's Q | Heterogeneity of genetic effects – pleiotropic effects | (25) |
| Steiger filtering | Hypothesized causal direction for each SNP | (28) |
| *Cis* Mendelian randomization | Pleiotropic effects of variants | (29) |

### Analyses including proxy cases of Alzheimer’s disease in UKB

The main analysis was conducted using summary data from phase one of the Alzheimer’s disease GWAS, which included only clinically diagnosed cases. To increase the statistical power of our analysis, we re-ran our analysis using summary data from phase three of Alzheimer’s GWAS, which includes proxy cases as defined in UKB (18). The decision to use phase three as a sensitivity analysis rather than as main analysis, was based on the following arguments: i. UK Biobank ‘cases’ included in this meta-analysis were not themselves diagnosed with Alzheimer’s disease (rather, it refers to the participants’ parent), ii. Cases are not doctor diagnosed but based on self-report and iii. The question asks about broad dementia, not specifically Alzheimer’s disease.

## RESULTS

### Selection of instruments & instrument strength

Out of the 41 cytokines we aimed to explore, 26 of them had at least one genetic variant available at the genome-wide significant threshold p<5×10^−08^ for use in our MR analyses. For an additional 8 cytokines, genetic variants were available at a more liberal threshold of p<5×10^−07^. For Alzheimer’s disease 29 genetic variants were available at the genome-wide significant threshold p<5×10^−08^, 26 of which were available in the cytokines GWAS. Additional information about the genetic instruments used in our analyses can be found in Table S1-S2. Notably, all selected instruments for cytokine concentrations and Alzheimer’s disease demonstrated an F-statistic larger than 10, indicating that weak instruments bias is unlikely to bias our results. Information on the number of instruments identified for each cytokine concentration, the threshold used for selecting instruments and the cumulative F-statistic per cytokine can be found in Table 2. Lastly, four genetic variants were identified as genetic instruments for more than one cytokine and Figure 1 illustrates the overlap. All these variants were included in our main analyses and their influence on our results was further explored in leave – one out analyses.

**Figure 1.**
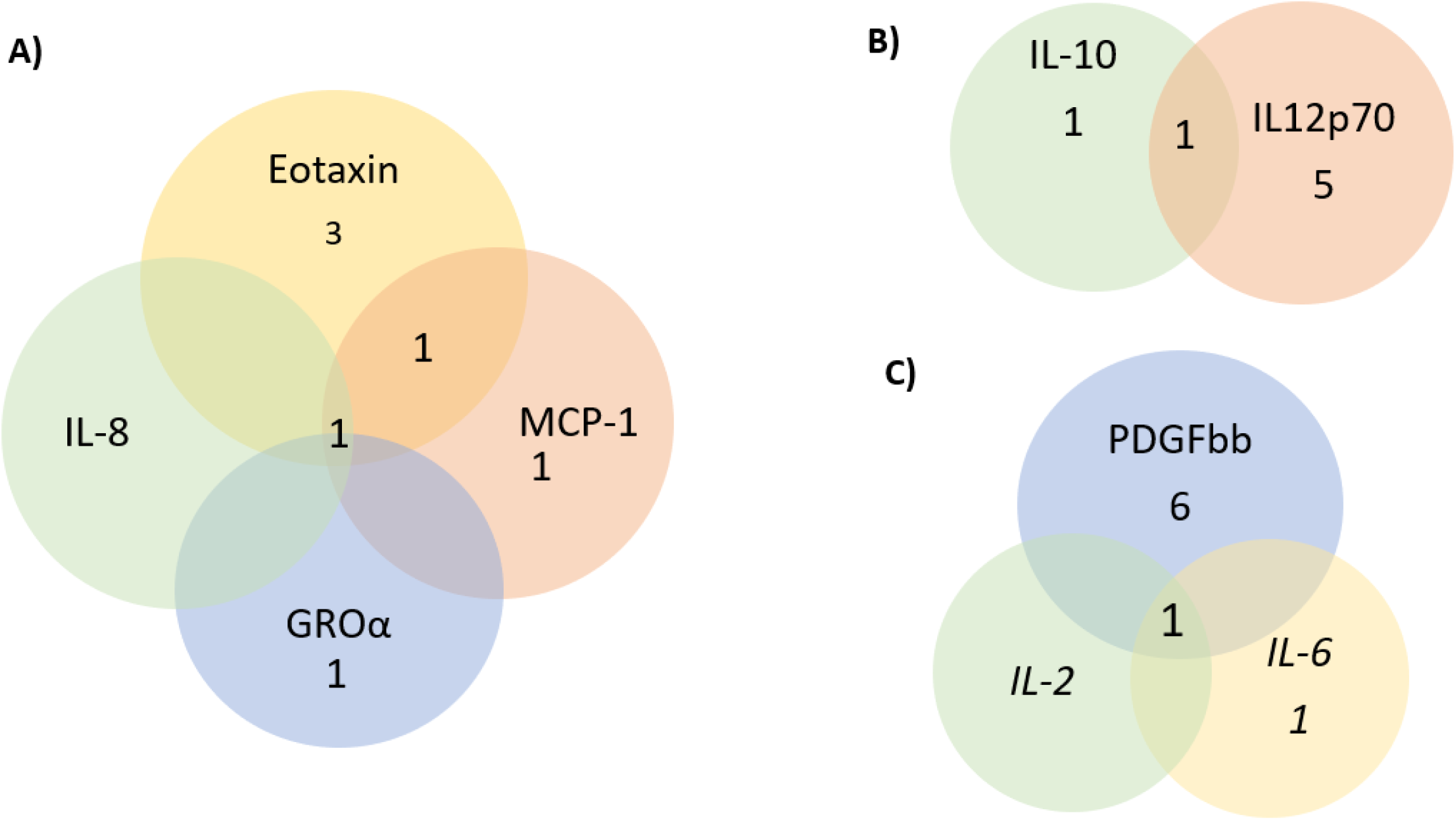
Graphical presentation of the number of genetic variants overlapping between cytokines

**Table 2.** Descriptive table of the number of participants for each circulating cytokine concentration, number of instruments identified for each cytokine, the threshold used for selecting instruments and the cumulative F-statistic

| **Cytokine abbreviation** | **Cytokine full name** | **N\*\*** | **No. of genetic instruments** | **Cumulative F-statistic** |
| --- | --- | --- | --- | --- |
| IL-6* | Interleukin-6 | 8,189 | 2 | 57.4 |
| IL-17 | Interleukin-17 | 7,760 | 1 | 38.9 |
| MCP1 (CCL2) | Monocyte chemotactic protein 1 | 8,293 | 4 | 422.5 |
| MIP1b (CCL4) | Macrophage inflammatory protein-1b | 8,243 | 25 | 2690.8 |
| GROa (CXCL1) | Growth regulated oncogene-alpha | 3,505 | 2 | 434.8 |
| IFNg* | Interferon gamma | 7,701 | 1 | 27.6 |
| IL-4* | Interleukin-4 | 8,124 | 2 | 51.5 |
| IL-10 | Interleukin-10 | 7,681 | 2 | 336.4 |
| IL-13 | Interleukin-13 | 3,557 | 2 | 322.9 |
| IL-7 | Interleukin-7 | 3,409 | 1 | 169.8 |
| IL-2ra | Interleukin-2 receptor alpha | 3,677 | 1 | 167.6 |
| IL12p70 | Interleukin-12p70 | 8,270 | 3 | 637.9 |
| IL-16 | Interleukin-16 | 3,483 | 3 | 203.6 |
| IL-18 | Interleukin-18 | 3,636 | 4 | 267.8 |
| CTACK (CCL27) | Cutaneous T-cell attracting | 3,631 | 4 | 261.9 |
| Eotaxin | Eotaxin | 8,153 | 5 | 425.3 |
| HGF | Hepatocyte growth factor | 8,292 | 2 | 98.0 |
| IP10 | Gamma-induced protein 10 | 3,685 | 1 | 31.1 |
| PDGFbb | Platelet derived growth factor BB | 8,293 | 7 | 549.1 |
| SCF | Stem cell factor | 8,290 | 2 | 80.4 |
| SCGFb | Stem cell growth factor-beta | 3,682 | 5 | 321.1 |
| TNFb | Tumour necrosis factor-beta | 1,559 | 2 | 130.8 |
| TRAIL | TNF-related apoptosis inducing ligand | 8,186 | 10 | 1484.7 |
| VEGF | Vascular endothelial growth factor | 7,118 | 9 | 1153.0 |
| MIG (CXCL9) | Monokine induced by interferon- gamma | 3,685 | 1 | 42.3 |
| RANTES (CCL5) | Regulated on Activation, Normal T Cell Expressed and Secreted | 3,421 | 1 | 29.9 |
| IL-1ra* | Interleukin -1 receptor alpha | 3,638 | 2 | 53.40 |
| IL-2* | Interleukin-2 | 3,475 | 1 | 28.2 |
| IL-5 | Interleukin-5 | 3,364 | 1 | 37.9 |
| IL-8* | Interleukin-8 | 3,526 | 1 | 25.8 |
| IL-9* | Interleukin-9 | 3,634 | 1 | 25.5 |
| MCP3* | Monocyte specific chemokine 3 | 843 | 1 | 25.6 |
| bNGF | beta nerve growth factor | 3,531 | 1 | 36.5 |
| MCSF | Macrophage colony-stimulating factor | 840 | 1 | 31.6 |

\* No genetic variants were available at the $p < 5 \times 10^{-08}$ threshold. Thus, genetic variants were identified using a more liberal threshold of $p < 5 \times 10^{-07}$ .
\*\* Number of participants included in the GWAS of each cytokine concentration.

### Causal effects of circulating cytokine concentrations on risk of Alzheimer’s disease

Overall, there was little evidence to support a causal effect of greater levels of circulating cytokines on risk of Alzheimer’s disease (Figure 2A-2C). Exception was CTACK (CCL27), where we observed weak evidence of a causal effect on Alzheimer’s disease (IVW: OR per 1 standard deviation (SD) increase =1.09 95%CI: 1.01 to 1.19, p=0.03). We also observed weak evidence of a causal effect of 1 SD increase in concentrations of MIP-1b (CCL4) (IVW: OR=1.04 95%CI: 0.99 to 1.09, p=0.08) and Eotaxin (IVW: OR=1.08 95%CI: 0.99 to 1.17, p =0.10) on risk of Alzheimer’s disease. Additionally, weak evidence was observed for an adverse effect of 1 SD increase in levels of GROa (CXCL1) (IVW: OR=1.04 95%CI: 0.99 to 1.10, p=0.15), MIG (CXCL9) (Wald Ratio: OR=1.17 95%CI: 0.97 to 1.41, p=0.10), IL-8 (Wald Ratio: OR=1.21 95%CI: 0.97 to 1.51, p=0.09) and IL-2 (Wald Ratio: OR=1.21 95%CI: 0.94 to 1.56, p=0.14).

**Figure 2A.**
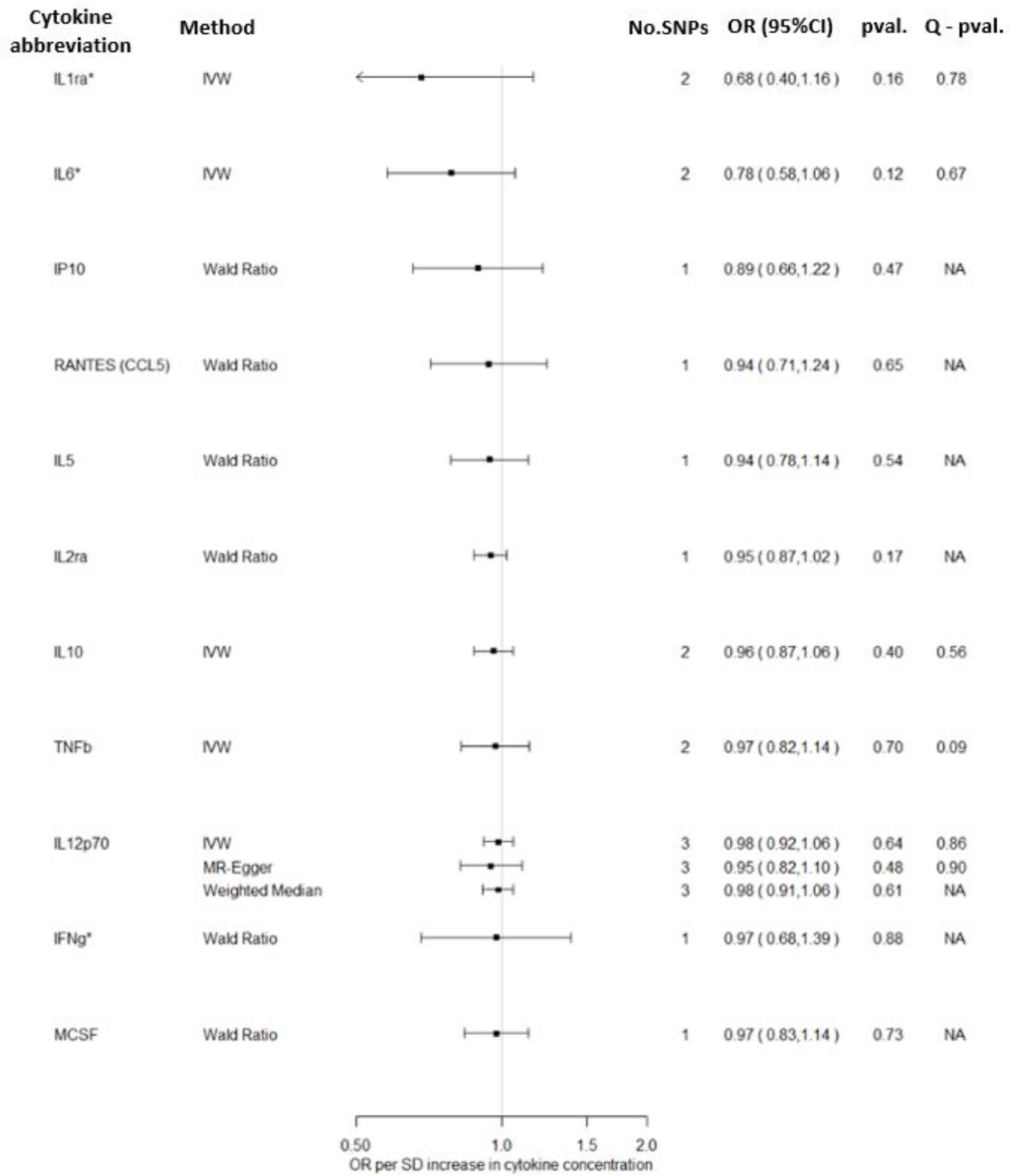
Total causal effect of genetically predicted cytokine concentrations on the risk of Alzheimer’s disease, as estimated by Wald Ratio, IVW, MR-Egger and Weighted median estimators.

**Figure 2B.**
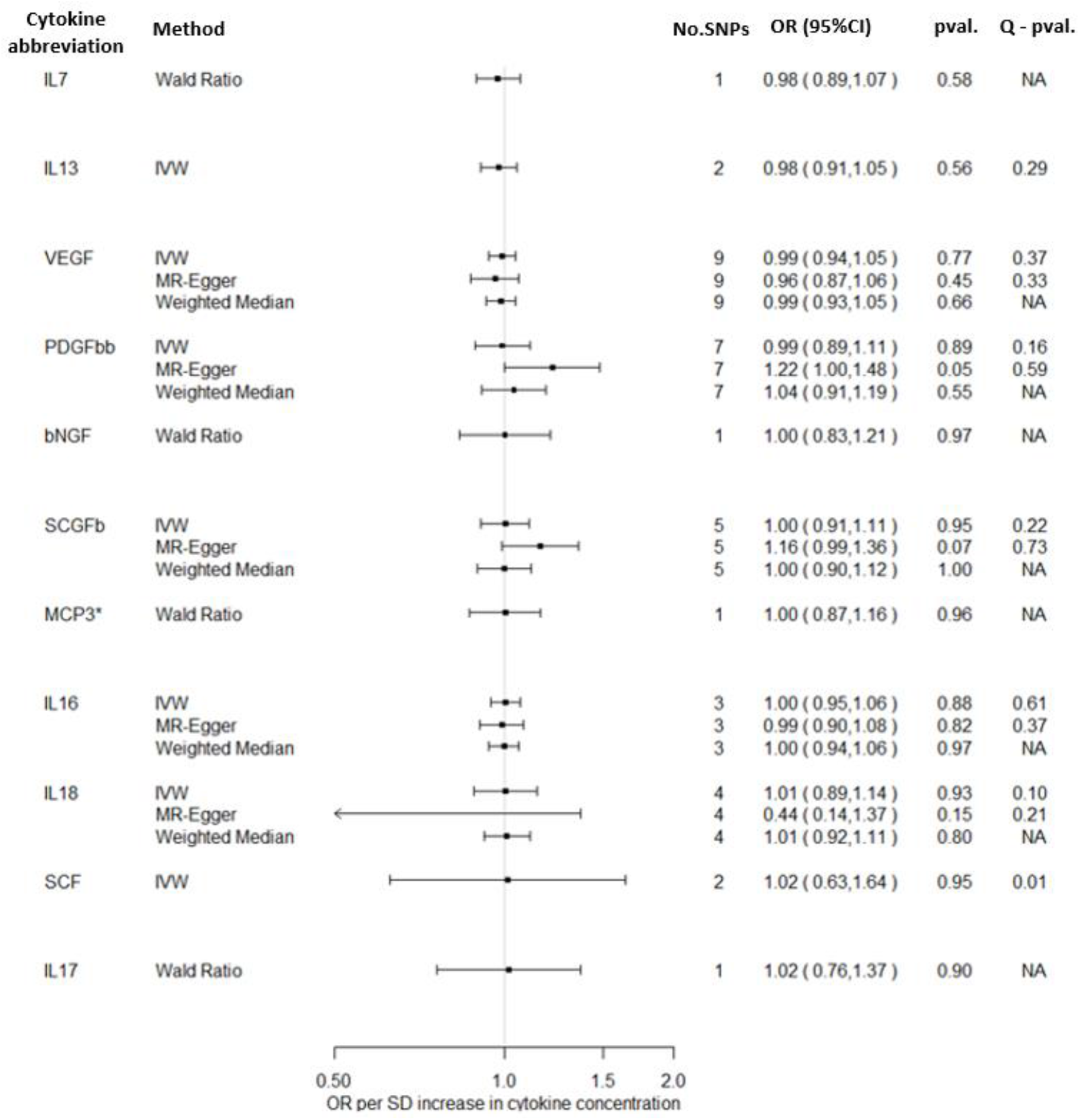
Total causal effect of genetically predicted cytokine concentrations on the risk of Alzheimer’s disease, as estimated by Wald Ratio, IVW, MR-Egger and Weighted median estimators.

**Figure 2C.**
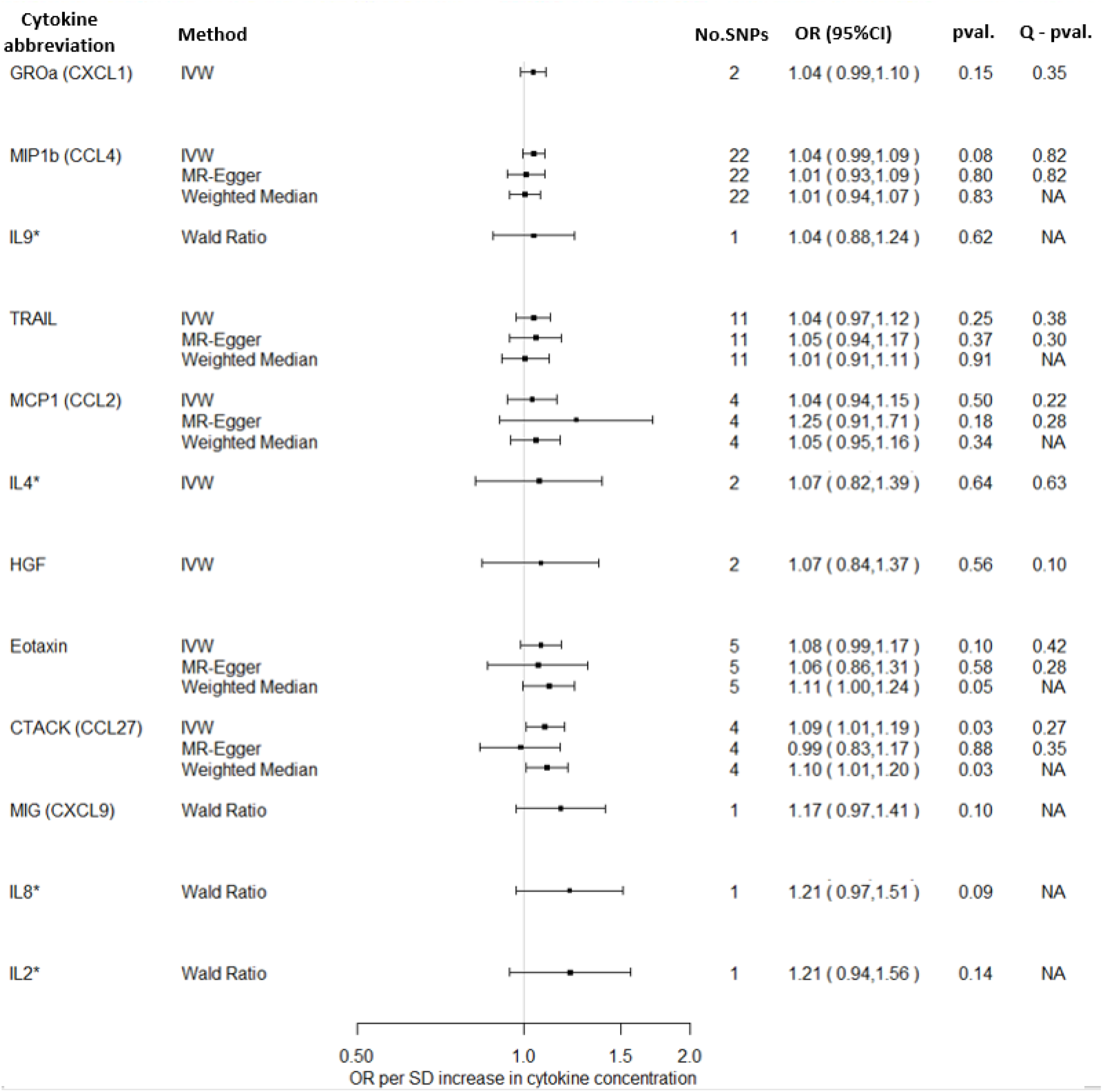
Total causal effect of genetically predicted cytokine concentrations on the risk of Alzheimer’s disease, as estimated by Wald Ratio, IVW, MR-Egger and Weighted median estimators.

### Sensitivity analyses

When more than three genetic instruments were available and MR-Egger and Weighted median could be estimated, we observed comparable results to the IVW. More specifically, for the cytokines we observed evidence for a causal effect of cytokines on Alzheimer’s disease, MR-Egger and Weighted median estimators yielded similar effect estimates to the IVW estimator (CTACK: MR-Egger slope: OR=0.99 95%CI: 0.83 to 1.17, p=0.88; Weighted median: OR=1.10 95%CI: 1.01 to 1.20, p =0.03; MIP-1b: MR-Egger slope: OR=1.02 95%CI: 0.95 to 1.11, p =0.58; Weighted median: OR=1.01 95%CI: 0.95 to 1.07, p =0.82; Eotaxin: MR-Egger slope: OR=1.06 95%CI: 0.86 to 1.31, p =0.58; Weighted median: OR=1.11 95%CI: 1.00 to 1.24, p =0.05).

There was no indication of heterogeneity as estimated by Cochran’s Q statistic and after iteratively removing genetic variants results remained virtually the same in the leave-one-out analysis (see Supplementary material Figure S1-S20). Steiger filtering suggested that none of the genetic variants used as instruments for cytokines explained more variation in the outcome, thus the larger sample size of the Alzheimer’s disease GWAS was unlikely to have affected our results and the direction of effects was correctly estimated.

When we restricted our analyses to variants located in closest proximity to the encoding gene of each cytokine, we obtained at least one *cis* variant for 11 cytokines that were initially included in our main analysis. Overall, results remained virtually the same, but with wider confidence intervals, as a smaller number of instruments was included (Table S3). For greater levels of the TRAIL cytokine we observed weak evidence of a detrimental effect (Wald Ratio: OR=1.23 95%CI: 0.97 to 1.55, p=0.07) on Alzheimer’s disease.

### Analyses including proxy cases of Alzheimer’s disease in UKB

Overall, there was limited evidence to suggest a causal effect of circulating cytokine concentrations on risk of Alzheimer’s disease (Supplementary Figures S21A-S21C). As the sample size increased significantly with the addition of proxy-cases, confidence intervals were narrower compared to analyses including only cases of Alzheimer’s disease. For IL-2, a detrimental causal effect on Alzheimer’s disease risk was observed (Wald Ratio: OR per 1 SD increase =1.03 95%CI: 1.00 to 1.06, p=0.04), although the effect estimate was much smaller in magnitude compared to analyses including only diagnosed cased of Alzheimer’s disease.

### Causal effects of genetic liability to Alzheimer’s disease on circulating cytokine concentrations

There was limited evidence to suggest a causal effect of genetic liability to Alzheimer’s disease on higher levels of circulating cytokine concentrations (Supplementary Figures 22A-22E). However, weak evidence was observed for a causal effect of genetic liability to Alzheimer’s disease on increased levels of circulating SCGFb (IVW: β= -0.06 95%CI: -0.12 to 0.01, p=0.08).

### Sensitivity analyses

Overall, MR-Egger and Weighted median yield similar results to IVW estimators. Additionally, there was no indication of heterogeneity as estimated by Cochran’s Q statistic and leave-one-out analysis did not indicate any genetic variants as influential.

### Analyses including proxy cases of Alzheimer’s disease in UKB

When we included the proxy-cases of Alzheimer’s disease in our analysis we observed limited evidence to suggest a causal effect of genetic liability to Alzheimer’s disease on circulating cytokine concentrations (Supplementary Figures S22A-S22E). However, the causal effect of genetic liability to Alzheimer’s disease on SCGFb concentrations was confirmed (IVW: β= -0.74 95%CI: -1.16 to -0.33, p<0.01). Notably, the estimated causal effect was greater in magnitude.

## DISCUSSION

Within a two-sample MR framework, we investigated the effect of 34 circulating cytokine concentrations on Alzheimer’s disease risk, and vice versa. We observed some evidence for a detrimental effect of greater levels of CTACK (CCL27), MIP-1b (CCL4), Eotaxin, GROa (CXCL1), MIG (CXCL9), IL-8 and IL-2 on the risk of Alzheimer’s disease. In the reverse direction, we observed some evidence to suggest a causal effect of genetic liability to Alzheimer’s disease only on levels of SCGFb.

Very few studies have examined the causal effects of circulating cytokine concentrations of Alzheimer’s disease risk. Previous two-sample MR studies have investigated the causal role of a few cytokines (TNF-a, IL-18, IL-1ra, IL-6) on risk of Alzheimer’s disease (13-16) and reported limited evidence to support a causal role. In our study we used a larger GWAS of Alzheimer’s disease, thus increasing statistical power to observe causal effects. Despite the increased statistical power, our results were in line with these findings suggesting there is little evidence of a causal effect of circulating cytokines on risk of Alzheimer’s disease. Notably, none of the previous two-sample MR studies have investigated the causal effect of genetic liability of Alzheimer’s disease on increased levels of cytokines.

Chemokines are cytokines which regulate immune cell migration and are thought to be mediators of the peripheral monocytes into the inflamed CNS (30), thus are hypothesized to be involved in the pathogenesis of Alzheimer’s disease (31). CTACK (CCL4), MIG (CXCL9), GROa (CXCL1), MIP-1b (CCL4), Eotaxin (CCL11) and IL-8 belong to the chemokine family and evidence suggest that they could potentially play a role in Alzheimer’s disease. More specifically, CTACK (CCL27) is a chemokine involved in the CNS as it is expressed in the cerebral cortex and limbic regions which are mainly affected in Alzheimer’s disease (32). Previous studies have observed higher levels of circulating CTACK (CCL27) in Alzheimer’s disease patients compared to healthy controls (33, 34). However, future research is required to better characterise the role of CTACK (CCL27) in Alzheimer’s disease aetiology.

MIG (CXCL9) is another chemokine that is considered to play a role in the interplay between neurons and glial cells, and binds onto the CXCR3 receptor which has been previously reported to be involved in the pathogenesis of various CNS conditions (e.g. multiple sclerosis, glioma, bipolar disorder) (35, 36). Regarding Alzheimer’s disease, a cross-sectional study reported substantially higher levels of circulating MIG (CXCL9) in patients with Alzheimer’s disease compared to non-cognitively impaired and mildly-cognitively impaired participants (34). Additionally, a recent case-control study demonstrated evidence for an association between higher levels of MIG and Alzheimer’s disease in a Mexican population (33).

A case-control study found that GROa (CXCL1) is overexpressed in the brains of 23 Alzheimer’s disease patients, with no prior diagnosis of immunological diseases, hypertension, cardiac disease or diabetes, compared to age-matched controls (37). This result is supported by animal studies, where CXCL1 was found to drive the hypermethylation of Tau in the primary cortical neurons of mice (38, 39). This is supporting to our findings and together, it suggests a plausible causal role of CXCL1 in the pathogenesis of Alzheimer’s disease.

Xia et al. reported that receptors of MIP-1b (CCL4) were present on microglia and subpopulation of reactive astrocytes and neurons in brains of patients with Alzheimer’s disease compared to controls, thus they could potentially play a role in the progression of Alzheimer’s disease through glial-glial and glial-neuronal interactions (40). Moreover, higher levels of MIP-1b has been associated with cognitive decline in patients with Alzheimer’s disease (41). Additionally, several studies have reported that expression of CCL4 is greater in brains of HIV-infected patients with dementia compared to HIV-infected patients without dementia, which indicates that CCL4 possibly regulates an inflammatory process that indirectly affects neurons (42-44).

Higher levels of Eotaxin were identified in the cerebrospinal fluid and serum of Alzheimer’s disease cases compared to healthy controls (41, 45, 46). In contrast, in two cohort studies, higher levels of Eotaxin in the plasma were not associated with Alzheimer’s disease progression (47, 48). However, all the studies are of small sample size and thus underpowered to identify associations.

Evidence regarding the role of IL-8 in the Alzheimer’s disease pathogenesis is contradicting. A case-control study observed elevated levels of IL-8 in the CSF of Alzheimer’s disease patients compared to non-demented controls (41), when another case-control study reported significantly lower levels in both serum and CSF of Alzheimer’s disease patients (49). Additionally, IL-8 plasma and CSF levels have been found to be associated with higher levels of p-tau and with higher levels of CSF Aβ1-42, which are hallmarks of Alzheimer’s disease (50). However, evidence stems from studies with limited number of participants and further research is required to elucidate the role of these chemokines in the pathogenesis and progression of Alzheimer’s disease.

### Strengths & limitations

The main strength of the present study is the use of two-sample MR which could aid in overcoming the drawbacks of traditional observational epidemiology (i.e. reverse causation, residual confounding), thus allowing the estimation of the effect size of the association between circulating cytokines and Alzheimer’s disease. Additionally, the plethora of cytokines which we could identify instruments for allowed us to explore causal associations between cytokine concentrations and Alzheimer’s disease. Following an agnostic approach could be beneficial in this setting as limited observational evidence exists for most of the studied cytokines. We also used the largest GWAS study of Alzheimer’s disease, which included 24,087 clinically diagnosed cases and thus, our study had statistical power to identify causal effects. The power to detect an odds ratio of 1.1 per 1 – SD increase in circulating cytokine concentration, with a = 0.05 and a coefficient of determination (R^2^) of 3% to 6%, was between 60% and 80%.

There are a few limitations of this study. As few of the genetic variants used in our MR analyses were used as instruments for more than one cytokine, we cannot exclude the possibility that our results were biased due to horizontal pleiotropic effects. We addressed this issue by estimating the total causal effects with alternative MR methods (MR-Egger and weighted median). Even though results remained virtually the same we could not completely exclude the possibility of pleiotropic effects as these methods are not reliable when a limited number of genetic variants is available, which is the case for many of the cytokines investigated.

Moreover, cytokines are complex in their activity as they act pleiotropically (i.e. single cytokine acts on several different cell types), are redundant in their activity (i.e. same process might be activated by multiple cytokines) and can act either synergistically or antagonistically (51). This complex activity of cytokines raises two main issues in our study. The first issue is the possibility that the observed causal effects are either confounded or mediated by one or more cytokines, and therefore not representing the direct causal effect of each cytokine (i.e. the causal effect after taking into account the causal effect of all other cytokines that might mediate or confound the observed total causal effect). A future direction in deciphering the direct causal effects of cytokines on risk of Alzheimer’s disease would be the employment of multivariable Mendelian randomization, which allows the estimation of direct causal effects (52, 53). MVMR could be used either by including cytokines which belong in the same structural family (i.e. chemokines, interleukins, haemopoietins, chemokines, interferons, TNF family, colony stimulating factors) in a single model or based on their anti/pro-inflammatory activity. However, we were not able to implement MVMR mostly due to the presence of correlation between the estimated genetic effects of the inflammatory markers, as they were obtained from the same participants. Additionally, the limited number of genetic instruments available for each cytokine would have resulted in weak instrument bias and would have limited the statistical power to identify direct effects in the MVMR framework (54). The second issue is that in our analyses we assume no gene-gene or gene-environment interactions and thus modelling the interplay between the multiple inflammatory markers examined is not feasible.

Lastly, observed causal effects might be the consequence of collider bias. Collider bias occurs when conditioning on a variable that is affected by both exposure and outcome and could lead to spurious associations (55, 56). In our study, the GWAS used to extract instruments for cytokine concentrations was adjusted for BMI, which is affected by both cytokines and Alzheimer’s disease, thus it could be considered as a collider. However, this is unlikely to affect our results as individuals on the cytokines GWAS were not selected based on their BMI measurement.

## CONCLUSION

In a two-sample MR framework we observed some evidence to support a causal role of cytokines in the pathogenesis of Alzheimer’s disease. More studies are needed to elucidate the specific mechanistic pathways that underlie this process. Better understanding of these processes could potentially lead to novel therapeutic targets for affected individuals.

## Data Availability

N/A

## Notes

**Funding:**This work was supported by a grant from the BRACE Alzheimer’s Disease charity (BR16/028). PP, ELA, and ES work in a unit that receives funding from the University of Bristol and the UK Medical Research Council (MC_UU_00011/1, MC_UU_00011/3). ELA is funded by an MRC Skills Development Award from the UK Medical Research Council (MR/P014437/1). LDH is funded by a Career Development Award from the UK Medical Research Council (MR/M020894/1). This publication is the work of the authors, and ELA, will serve as a guarantor for the contents of this paper.

### Competing Interest Statement

The authors have declared no competing interest.

### Funding Statement

This work was supported by a grant from the BRACE Alzheimers Disease charity (BR16/028). PP, ELA, and ES work in a unit that receives funding from the University of Bristol and the UK Medical Research Council (MC_UU_00011/1, MC_UU_00011/3). ELA is funded by an MRC Skills Development Award from the UK Medical Research Council (MR/P014437/1). LDH is funded by a Career Development Award from the UK Medical Research Council (MR/M020894/1). This publication is the work of the authors, and ELA, will serve as a guarantor for the contents of this paper.

