## Supplementary material for "Causal effects of circulating cytokine concentrations on risk of Alzheimer’s disease: A bidirectional two-sample Mendelian randomization study"

#### Supplementary Note 1. Harmonization of alleles

Before estimating total causal effects, we harmonised our datasets so that the effect estimates of each exposure were coded to express the effect estimate per increasing allele, so that outcome alleles correspond to the same alleles as in the exposure. For genetic variants which are ambiguous (i.e. the forward strand is not clear) the effect allele frequencies of both exposure and outcome are required to employ the harmonization. However, in our analysis allele frequencies were not available in the GWAS of cytokines concentrations and therefore alignment of alleles between exposure and outcome was not feasible in a few cases, resulting in the exclusion of those ambiguous genetic variants due to being "palindromic".

#### Supplementary Note 2. Sensitivity Analyses

As the validity of MR depends largely on the strength of the genetic instruments, we used the F-statistic to evaluate whether weak instrument bias could have affected our results (1). An F-statistic smaller than 10 indicates that weak instrument bias may be present and causal effect estimates could be biased. Secondly, the estimated total causal effects obtained from IVW method were compared with those obtained from MR-Egger regression and weighted median estimators. Unlike IVW, MR-Egger regression allows for an unconstrained intercept term and provides a robust causal effect estimate, after adjusting for horizontal pleiotropy (2). The weighted median estimator serves as an unbiased causal effect estimate when up to 50% of the instruments are invalid, by estimating the causal effect as the median of the weighted ratio estimates (3). When results from the above methods agree in direction and magnitude, we consider them more likely to be valid. Additionally, the influence of each genetic variant on the outcome was explored by conducting a leave-one out analysis, where genetic variants were systematically removed and causal effects of the remaining SNPs on the outcome were re-estimated (4). In cases where less than three genetic variants were used as instruments, estimation of the MR-Egger and weight median and leave-one out analysis were not feasible. Cochran's Q statistic was used to assess if the causal estimates of all genetic variants within a single MR analysis were comparable.

Substantial heterogeneity is an indication that instruments may not be valid (2). We also applied Steiger to test that the hypothesised causal direction was correct for each SNP (i.e. that that the instruments explain more variation in the exposure than the outcome) (5). Lastly, to further explore the issue of pleiotropic effects of genetic variants, we repeated analyses using only cis-variants (i.e. variants located in the closest proximity to the encoding gene of each cytokine) which are less likely to violate the 'horizontal pleiotropy assumption' than variants who are located more distantly (6). Further information about the procedure followed to extract cis-variants and perform cis-MR can be found in Supplementary Note 3.

**Supplementary Note 3. Cis-Mendelian randomization**

We defined as *cis* variants all genetic variants located within a 500kb window at either side of the gene that encodes each cytokine. When *cis* variants were available for a cytokine, we further selected independent genome-wide significant variants ($r^2<0.01$ within a 10,000 kb window, $p<5\times10^{-08}$) and corresponding SD-scaled effect sizes and standard errors were extracted from the publicly available datasets (7). After finalizing the list of *cis* variants for each cytokine, we extracted the corresponding log odds and standards errors from the Alzheimer's disease GWAS. Lastly, we performed harmonisation of alleles and estimated the total causal effects of each cytokine concentration on the risk of Alzheimer's disease using the same statistical analysis as in our main analysis (i.e. Wald ratio, IVW, MR-Egger, Weighted Median). More information about the location of each encoding gene and the number of resulting *cis* variants can be found in Table S3

### TABLES

**Table S1.** Characteristics of genetic variants associated with each circulating cytokine concentration.

| Cytokine abbreviation | SNP ID | Chromosome | trans/cis | Effect/ Ref allele | Beta (SE) | P-value |
|---|---|---|---|---|---|---|
| IL-6* | **rs13412535** | 2 | trans | G/A | 0.1164 (0.0215) | 7.34E-08 |
| | rs72831623 | 17 | trans | A/G | 0.1973 (0.0372) | 1.08E-07 |
| IL-17 | rs1530455 | 3 | trans | T/C | 0.1080 (0.0173) | 4.87E-10 |
| MCP-1 (CCL2) | **rs12075** | 1 | trans | A/G | 0.2185 (0.0155) | 1.44E-44 |
| | rs2036297 | 3 | trans | A/G | 0.1190 (0.0160) | 1.09E-13 |
| | **rs2228467** | 3 | trans | C/T | 0.2637 (0.0291) | 9.19E-20 |
| | rs7632755 | 3 | trans | A/G | 0.2938 (0.0316) | 1.18E-20 |
| MIP1b (CCL4) | rs113010081 | 3 | trans | C/T | 0.5954 (0.0236) | 3.85E-140 |
| | rs2673050 | 3 | trans | T/G | 0.1314 (0.0161) | 3.14E-16 |
| | rs4683315 | 3 | trans | G/A | 0.1347 (0.0236) | 8.97E-09 |
| | rs73074316 | 3 | trans | G/A | 0.1436 (0.0216) | 2.80E-11 |
| | rs79091774 | 3 | trans | C/A | 0.4606 (0.0751) | 8.83E-10 |
| | rs10491120 | 17 | trans | A/G | 0.3001 (0.0318) | 5.15E-21 |
| | rs111942332 | 17 | trans | G/T | 0.4727 (0.0573) | 1.70E-16 |
| | rs113877493 | 17 | cis | C/T | 0.6124 (0.0218) | 1.62E-173 |
| | rs117453826 | 17 | trans | G/A | 0.5774 (0.0593) | 5.07E-22 |
| | rs117620244 | 17 | trans | C/T | 0.3528 (0.0495) | 1.87E-12 |
| | rs117715247 | 17 | cis | G/A | 0.3510 (0.0590) | 3.09E-09 |
| | rs17693183 | 17 | trans | G/A | 0.5795 (0.0795) | 8.93E-13 |
| | rs2190980 | 17 | trans | G/A | 0.1067 (0.0167) | 1.43E-10 |
| | rs4795162 | 17 | Trans | A/G | 0.1261 (0.0158) | 1.14E-15 |
| | rs4796072 | 17 | trans | G/T | 0.1149 (0.0177) | 8.12E-11 |
| | rs60516659 | 17 | cis | A/G | 0.2691 (0.0248) | 3.64E-27 |
| | rs6505501 | 17 | trans | C/T | 0.1556 (0.0191) | 3.71E-16 |
| | rs7221878 | 17 | trans | C/T | 0.3045 (0.0463) | 7.37E-11 |
| | rs76842834 | 17 | trans | C/T | 0.4206 (0.0472) | 7.33E-19 |
| | rs76960253 | 17 | cis | T/C | 0.5233 (0.0585) | 5.45E-19 |
| | rs80007108 | 17 | trans | A/C | 0.2259 (0.0310) | 2.73E-13 |
| | rs9330240 | 17 | trans | C/T | 0.4745 (0.0460) | 5.84E-25 |
| GROa (CXCL1) | **rs12075** | 1 | trans | A/G | 0.3751 (0.0237) | 1.24E-55 |
| | rs508977 | 4 | cis | G/T | 0.3802 (0.0280) | 7.56E-42 |
| IFNg* | rs78296352 | 1 | trans | T/G | 0.3430 (0.0652) | 1.38E-07 |
| IL-4* | rs17713451 | 7 | trans | A/G | 0.1274 (0.0253) | 4.97E-07 |
| | rs10512267 | 9 | trans | C/T | 0.0824 (0.0161) | 2.94E-07 |
| IL-10 | rs282258 | 2 | trans | T/C | 0.0992 (0.0162) | 1.00E-09 |
| | **rs4349809** | 6 | trans | T/G | 0.2853 (0.0165) | 5.77E-67 |
| IL-13 | rs75438658 | 6 | trans | C/T | 0.3430 (0.0625) | 4.12E-08 |
| | rs9472168 | 6 | trans | A/G | 0.4244 (0.0248) | 1.08E-65 |
| IL-7 | rs4320361 | 6 | trans | G/T | 0.3245 (0.0249) | 6.87E-39 |
| IL-2ra | rs12722497 | 10 | cis | A/C | 0.6279 (0.0485) | 1.57E-38 |
| IL12p70 | rs12199215 | 6 | trans | T/C | 0.1278 (0.0192) | 5.11E-11 |
| | **rs4349809** | 6 | trans | T/G | 0.3777 (0.0159) | 2.56E-124 |
| | rs7754905 | 6 | trans | G/A | 0.1029 (0.0190) | 4.28E-08 |
| IL-16 | rs4253283 | 4 | trans | T/C | 0.1460 (0.0262) | 1.75E-08 |
| | rs1801020 | 5 | trans | A/G | 0.1733 (0.0272) | 4.53E-10 |
| | rs4778636 | 15 | cis | G/A | 0.7272 (0.0633) | 1.11E-30 |
| IL-18 | rs385076 | 2 | trans | C/T | 0.2432 (0.0248) | 1.66E-22 |

| | | | | | | |
|---|---|---|---|---|---|---|
| | rs116656892 | 5 | trans | T/C | 0.5298 (0.0925) | 1.05E-08 |
| | rs17229943 | 5 | trans | C/A | 0.3120 (0.0463) | 1.62E-11 |
| | rs71478720 | 11 | cis | C/T | 0.2669 (0.0276) | 3.71E-22 |
| CTACK (CCL27) | rs2070074 | 9 | cis | A/G | 0.4467 (0.0374) | 1.79E-32 |
| | rs58704839 | 9 | cis | A/G | 0.1785 (0.0284) | 3.29E-10 |
| | rs55764737 | 15 | trans | T/C | 0.5313 (0.0972) | 4.62E-08 |
| | rs135564 | 22 | trans | G/A | 0.1893 (0.0268) | 2.43E-12 |
| Eotaxin | **rs12075** | 1 | trans | A/G | 0.1671 (0.0156) | 1.33E-26 |
| | **rs2228467** | 3 | trans | C/T | 0.4163 (0.0292) | 2.27E-46 |
| | rs3091309 | 3 | trans | A/G | 0.1283 (0.0203) | 3.63E-10 |
| | rs342511 | 3 | trans | A/G | 0.0927 (0.0157) | 3.60E-09 |
| | rs2024050 | 7 | cis | A/G | 0.1728 (0.0303) | 1.10E-08 |
| HGF | rs3748034 | 4 | trans | T/G | 0.1495 (0.0234) | 1.81E-10 |
| | rs5745687 | 7 | cis | C/T | 0.3072 (0.0406) | 2.75E-14 |
| IP10 | rs113831257 | 4 | cis | A/G | 0.3592 (0.0644) | 2.53E-08 |
| PDGFbb | rs12990266 | 2 | trans | A/G | 0.2363 (0.0342) | 3.18E-12 |
| | rs13024765 | 2 | trans | C/T | 0.1014 (0.0158) | 1.14E-10 |
| | **rs13412535** | 2 | trans | A/G | 0.3352 (0.0214) | 2.46E-55 |
| | rs2324229 | 6 | trans | T/C | 0.0894 (0.0161) | 3.48E-08 |
| | rs28406863 | 15 | trans | G/T | 0.2089 (0.0382) | 4.78E-08 |
| | rs4965869 | 15 | trans | T/C | 0.1840 (0.0181) | 5.66E-24 |
| | rs9806745 | 15 | trans | A/C | 0.1162 (0.0163) | 1.10E-12 |
| SCF | rs1557570 | 1 | trans | T/G | 0.1186 (0.0170) | 2.74E-12 |
| | rs4841899 | 9 | trans | C/T | 0.1004 (0.0178) | 1.78E-08 |
| SCGFb | rs4656185 | 1 | trans | A/G | 0.2050 (0.0256) | 1.16E-15 |
| | rs17876031 | 5 | trans | G/A | 0.1514 (0.0255) | 2.25E-09 |
| | rs117716477 | 12 | trans | A/C | 0.8384 (0.0841) | 1.34E-23 |
| | rs73185877 | 12 | trans | A/G | 0.5249 (0.0711) | 1.18E-13 |
| | rs116924815 | 19 | trans | T/C | 0.6079 (0.0738) | 1.74E-16 |
| TNF-b | rs116196280 | 1 | trans | T/G | 0.7179 (0.1006) | 4.98E-13 |
| | rs78296352 | 1 | trans | T/G | 1.2215 (0.1366) | 4.76E-21 |
| TRAIL | rs3136596 | 3 | cis | A/G | 0.1147 (0.0209) | 3.65E-08 |
| | rs79287178 | 3 | cis | G/A | 0.4317 (0.0421) | 9.12E-25 |
| | rs11081739 | 18 | trans | A/G | 0.1411 (0.0202) | 3.34E-12 |
| | rs193112415 | 18 | trans | C/T | 1.0421 (0.0623) | 2.15E-62 |
| | rs57396456 | 18 | trans | C/T | 0.5626 (0.0518) | 1.25E-27 |
| | rs62093514 | 18 | trans | T/C | 1.0618 (0.0552) | 6.86E-82 |
| | rs62093947 | 18 | trans | C/T | 0.7596 (0.046) | 3.31E-61 |
| | rs74778900 | 18 | trans | T/C | 0.5906 (0.0532) | 2.59E-28 |
| | rs77451439 | 18 | trans | G/A | 0.4322 (0.0369) | 1.21E-31 |
| | rs9952273 | 18 | trans | T/C | 0.8640 (0.0499) | 3.86E-69 |
| VEGF | rs41282660 | 6 | cis | G/A | 0.1613 (0.0263) | 1.33E-09 |
| | rs4507572 | 6 | cis | T/C | 0.1007 (0.0171) | 3.34E-09 |
| | rs67798973 | 6 | cis | A/G | 0.1389 (0.0175) | 1.29E-15 |
| | rs6920532 | 6 | cis | T/C | 0.1803 (0.0267) | 8.68E-12 |
| | rs6921438 | 6 | cis | G/A | 0.4900 (0.0175) | 2.09E-171 |
| | rs74675876 | 6 | cis | C/T | 0.2822 (0.0366) | 7.62E-15 |
| | rs9381249 | 6 | cis | C/T | 0.2482 (0.0397) | 3.09E-10 |
| | rs9472183 | 6 | cis | G/A | 0.1282 (0.017) | 5.19E-14 |
| | rs34881325 | 9 | trans | C/T | 0.1082 (0.0189) | 1.04E-08 |
| MIG (CXCL9) | rs55876513 | 4 | cis | T/G | 0.1660 (0.0255) | 8.23E-11 |
| RANTES (CCL5) | rs74472919 | 13 | trans | T/C | 0.3313 (0.0605) | 3.97E-08 |
| IL-1ra* | rs146151667 | 2 | trans | A/G | 0.5072 (0.0978) | 3.36E-07 |
| | rs35803309 | 7 | trans | A/AT | 0.2080 (0.0404) | 2.36E-07 |

| | | | | | | |
|---|---|---|---|---|---|---|
| IL-2* | **rs13412535** | 2 | trans | A/G | 0.1764 (0.0332) | 1.18E-07 |
| IL-5 | rs7767396 | 6 | trans | A/G | 0.1515 (0.0246) | 7.69E-10 |
| IL-8* | **rs12075** | 1 | trans | A/G | 0.1200 (0.0236) | 3.88E-07 |
| IL-9* | rs76963786 | 12 | trans | C/T | 0.2865 (0.0557) | 4.50E-07 |
| MCP3* | rs10892381 | 11 | trans | T/C | 0.2412 (0.0476) | 3.56E-07 |
| bNGF | rs28637706 | 19 | trans | G/T | 0.1589(0.0263) | 1.42E-09 |
| MCSF | rs56367447 | 8 | trans | C/T | 0.4967(0.0883) | 1.72E-08 |

* No genetic variants were available on $p < 5\times10^{-08}$ threshold. Thus, genetic variants were identified using a more liberal threshold of $p < 5\times10^{-07}$.

**Table S2.** Characteristics of genetic variants associated with genetic liability to Alzheimer's disease (Phase1) (8)

| SNP ID | Chromosome | Effect/ Ref allele | Beta (SE) | P-value |
|---|---|---|---|---|
| rs2093760 | 1 | A/G | 0.1479 (0.0162) | 1.14E-19 |
| rs13004848 | 2 | C/T | 0.1202 (0.0205) | 4.62E-09 |
| rs6733839 | 2 | T/C | 0.1845 (0.0161) | 2.60E-30 |
| rs7657553 | 4 | A/G | 0.0884 (0. 0159) | 2.92E-08 |
| rs9272561 | 6 | G/A | 0.1367 (0. 0222) | 8.75E-10 |
| rs9381563 | 6 | C/T | 0.0922 (0. 0154) | 2.16E-09 |
| rs11763230 | 7 | C/T | 0.1269 (0. 0185) | 7.25E-12 |
| rs1859788 | 7 | G/A | 0.0948 (0. 0147) | 1.25E-10 |
| rs11787077 | 8 | C/T | 0.1399 (0. 0140) | 2.09E-23 |
| rs28834970 | 8 | C/T | 0.0850 (0. 0149) | 1.46E-08 |
| rs10792832 | 11 | G/A | 0.1290 (0. 0149) | 5.90E-18 |
| rs11218343 | 11 | T/C | 0.2577 (0. 0381) | 1.51E-11 |
| rs7935829 | 11 | A/G | 0.1100 (0. 0134) | 3.55E-16 |
| rs12590654 | 14 | G/A | 0.0926 (0. 0162) | 1.09E-08 |
| rs8093731 | 18 | C/T | 0.6136 (0. 1123) | 4.66E-08 |
| rs10407439 | 19 | G/A | 0.2455 (0. 0180) | 1.00E-200 |
| rs11666329 | 19 | A/G | 0.2274 (0. 0157) | 1.00E-200 |
| rs117310449 | 19 | T/C | 1.0823 (0. 0792) | 1.00E-200 |
| rs2376866 | 19 | T/C | 0.1637 (0. 0208) | 4.51E-15 |
| rs3865444 | 19 | C/A | 0.0898 (0. 0160) | 2.17E-08 |
| rs4147929 | 19 | A/G | 0.1219 (0. 0205) | 2.93E-09 |
| rs484195 | 19 | G/A | 0.3824 (0. 0205) | 1.00E-200 |
| rs55923289 | 19 | C/T | 0.3358 (0. 0234) | 1.00E-200 |
| rs595290 | 19 | C/T | 0.1564 (0. 0197) | 2.77E-15 |
| rs7412 | 19 | C/T | 0.4254 (0. 0359) | 2.63E-32 |
| rs899087 | 19 | T/C | 0.1952 (0. 0323) | 1.65E-09 |

**Table S3.** Causal effects of genetically determined cytokine concentrations on the risk of Alzheimer's disease, as obtained from the cis-MR analysis.

| Cytokine abbreviation | Gene location | No. SNPs | Wald Ratio OR (95% CI) | IVW OR (95% CI) | MR-Egger slope OR (95% CI) | Weighted Median OR (95%CI) |
|---|---|---|---|---|---|---|
| IL2ra | Chr 10: 6,052,652 - 6,104,333 | 1 | 0.94 (0.87 to 1.02) p=0.166 | N/A | N/A | N/A |
| VEGF | VEGF A: Chr6: 43,737,921-43,754,224<br>VEGFB: Chr11: 64,002,010-64,006,259<br>VEGFC: Chr4: 177,604,689-177,713,895 | 8 | N/A | 0.99 (0.93 to 1.05) p=0.796 | 0.96 (0.86 to 1.06) p=0.460 | 0.98 (0.93 to 1.04) p= 0.666 |
| IL-16 | Chr 15: 81,451,916-81,606,399 | 1 | 0.99 (0.93 to 1.06) p=0.930 | N/A | N/A | N/A |
| IL-18 | Chr 11: 112,013,974-112,034,840 | 1 | 0.97 (0.86 to 1.10) p=0.720 | N/A | N/A | N/A |
| GROa (CXCL1) | Chr 4: 74,735,110-74,737,025 | 1 | 1.00 (0.92 to 1.09) p=0.850 | N/A | N/A | N/A |
| MIP1b (CCL4) | Chr17: 34,430,983-34,433,014 | 6 | N/A | 1.00 (0.90 to 1.11) p=0.902 | 1.02 (0.75 to 1.39) p=0.857 | 0.96 (0.84 to 1.10) p=0.605 |
| TRAIL | Chr 3: 172,223,298-172,241,265 | 2 | N/A | 1.23 (0.97 to 1.55) p=0.07 | N/A | N/A |
| HGF | Chr 7: 81,328,326-81,399,754 | 1 | 0.99 (0.83 to 1.18) p=0.944 | N/A | N/A | N/A |
| Eotaxin | Chr 7: 75,440,983-75,452,674 | 1 | 1.24 (0.92 to 1.66) p=0.142 | N/A | N/A | N/A |
| CTACK (CCL27) | Chr 9: 34,661,877-34,664,045 | 2 | N/A | 1.06 (0.91 to 1.24) p=0.412 | N/A | N/A |
| MIG (CXCL9) | Chr 4: 76,922,428-76,928,662 | 1 | 1.16 (0.96 to 1.40) p=0.103 | N/A | N/A | N/A |

**FIGURES**

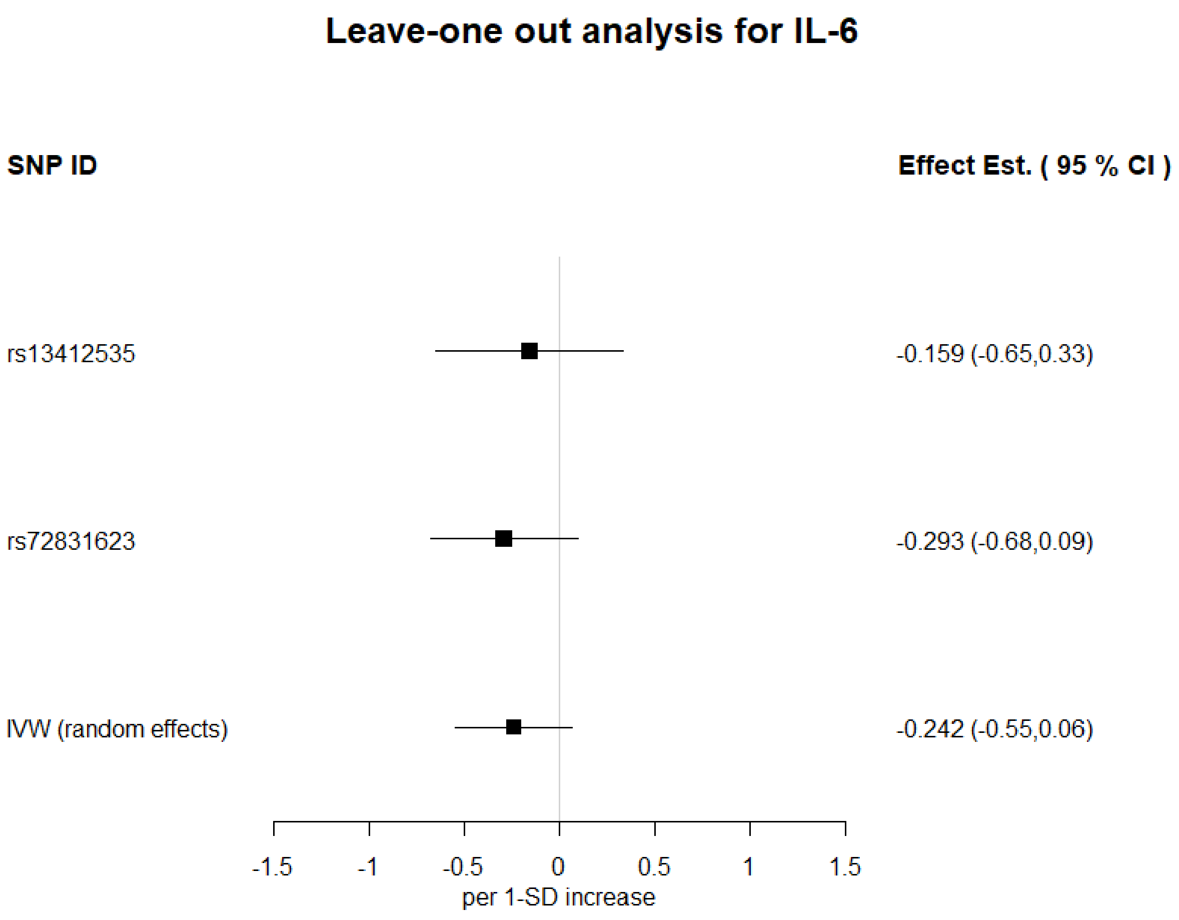


**Figure S1.** Leave-one out analyses of 1-SD increase in IL-6 on Alzheimer's disease, using IVW. Estimates shown as log odds and 95%CI

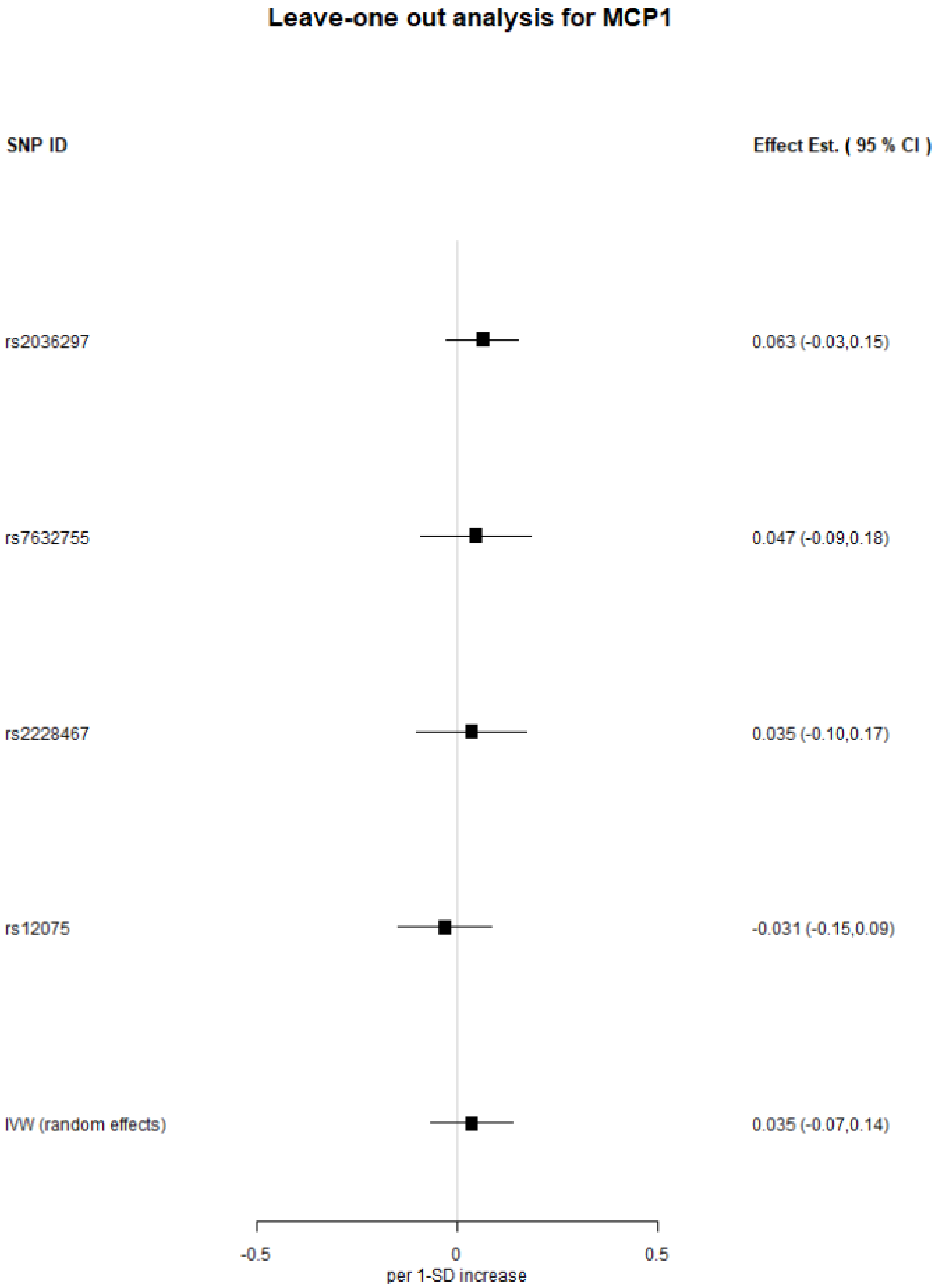


**Figure S2.** Leave-one out analyses of 1-SD increase in MCP1 on Alzheimer's disease, using IVW. Estimates shown as log odds and 95%CI

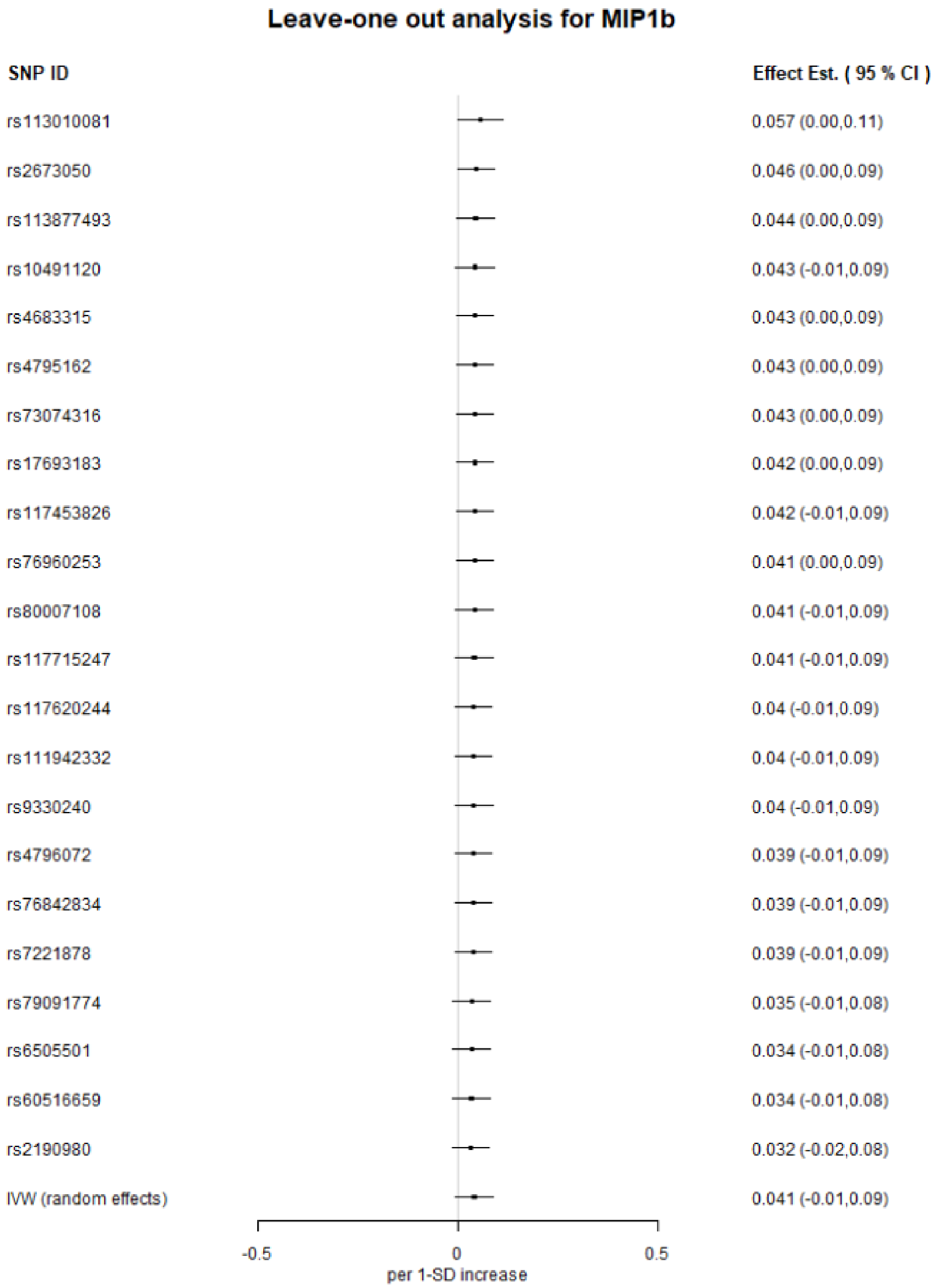


**Figure S3.** Leave-one out analyses of 1-SD increase in MIP1b on Alzheimer's disease, using IVW. Estimates shown as log odds and 95%CI

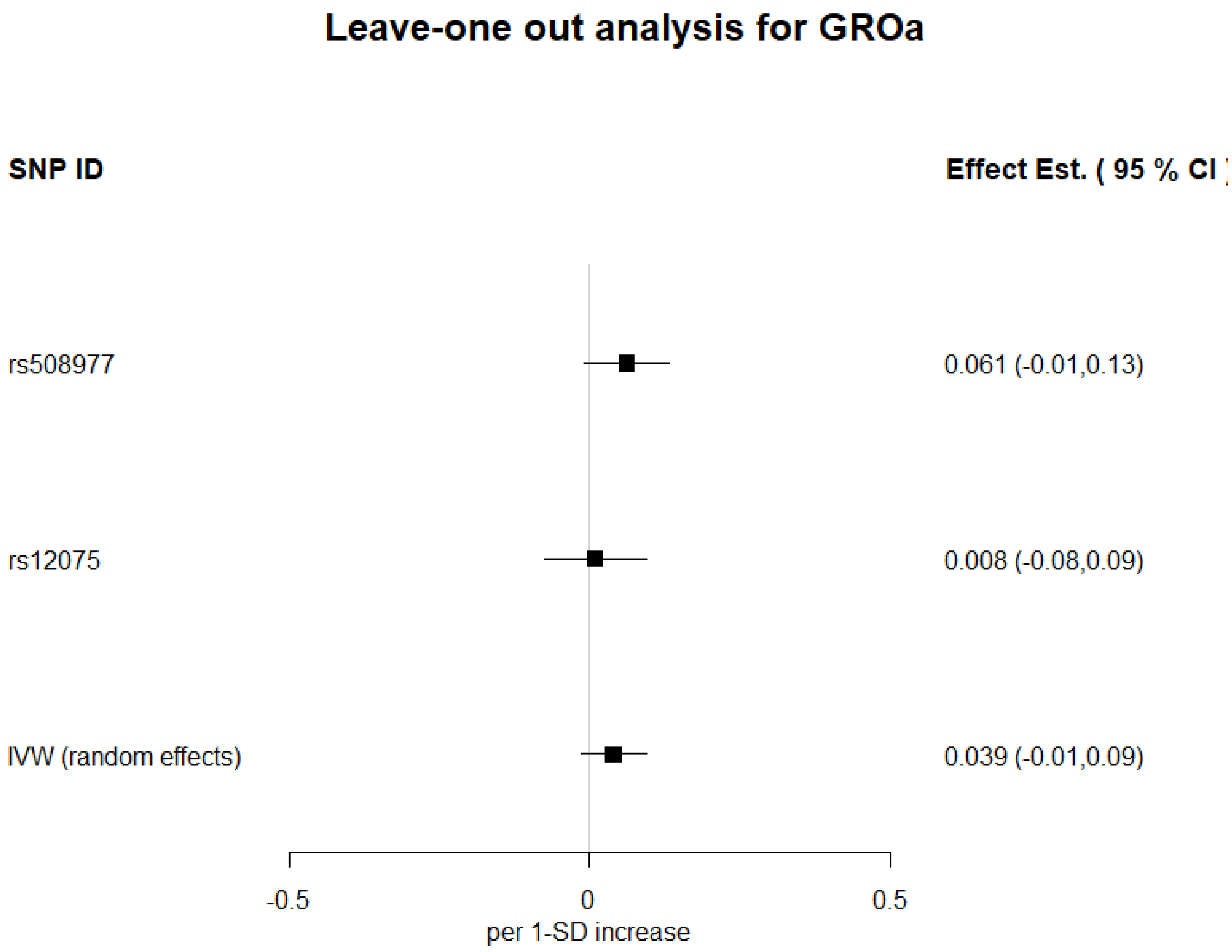


**Figure S4.** Leave-one out analyses of 1-SD increase in GROa on Alzheimer's disease, using IVW. Estimates shown as log odds and 95%CI

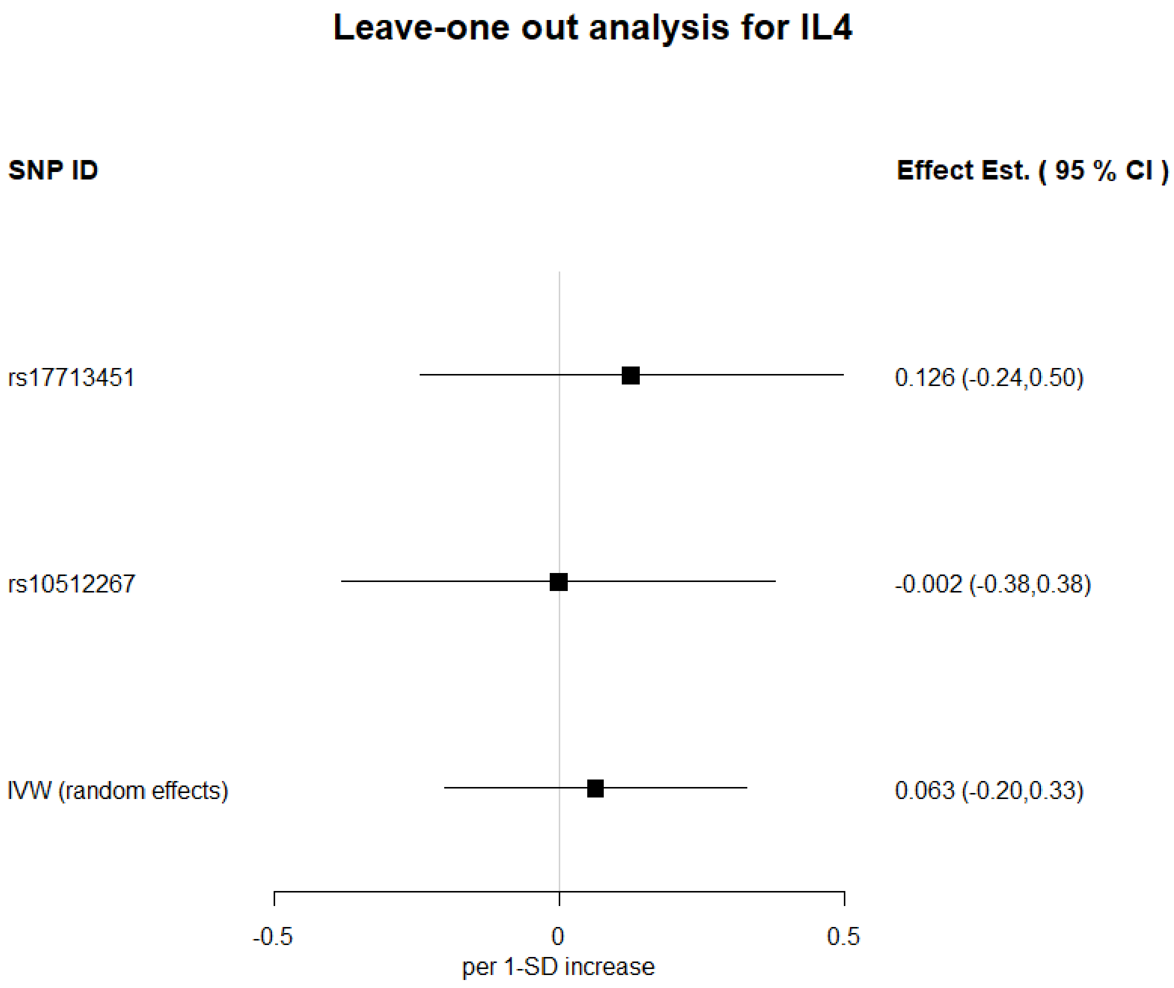


**Figure S5.** Leave-one out analyses of 1-SD increase in IL-4 on Alzheimer's disease, using IVW. Estimates shown as log odds and 95%CI

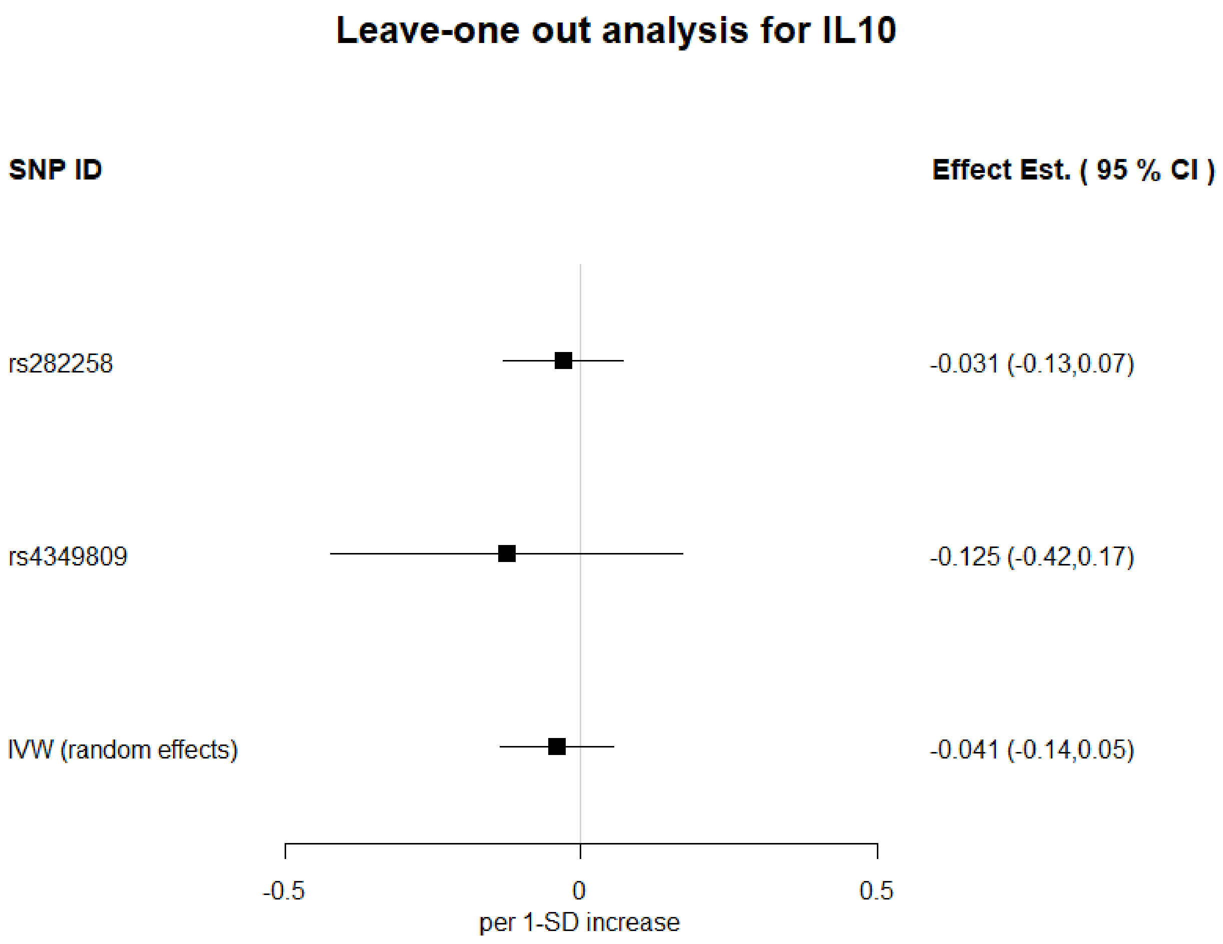


**Figure S6.** Leave-one out analyses of 1-SD increase in IL-10 on Alzheimer's disease, using IVW. Estimates shown as log odds and 95%CI

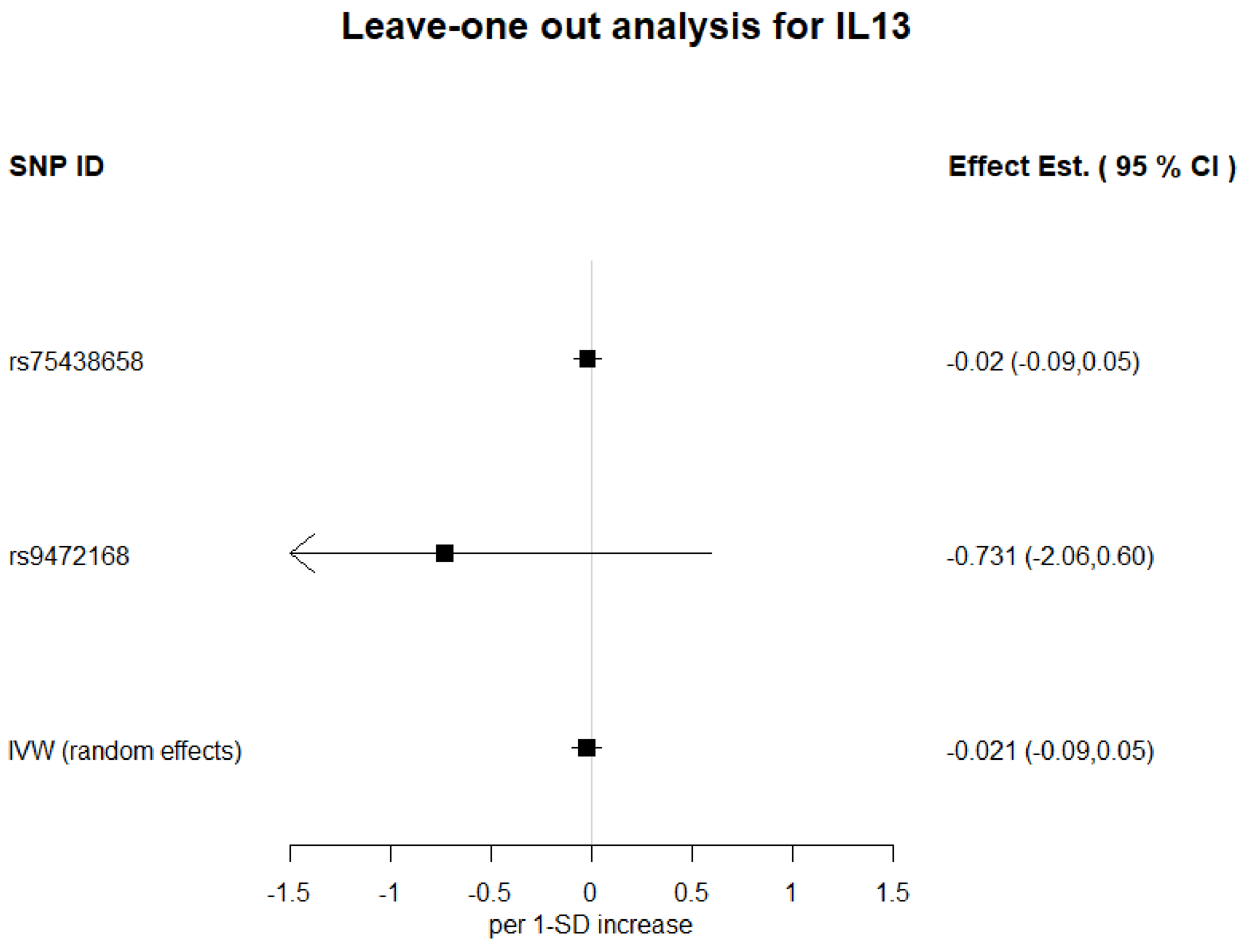


**Figure S7.** Leave-one out analyses of 1-SD increase in IL-13 on Alzheimer's disease, using IVW. Estimates shown as log odds and 95%CI

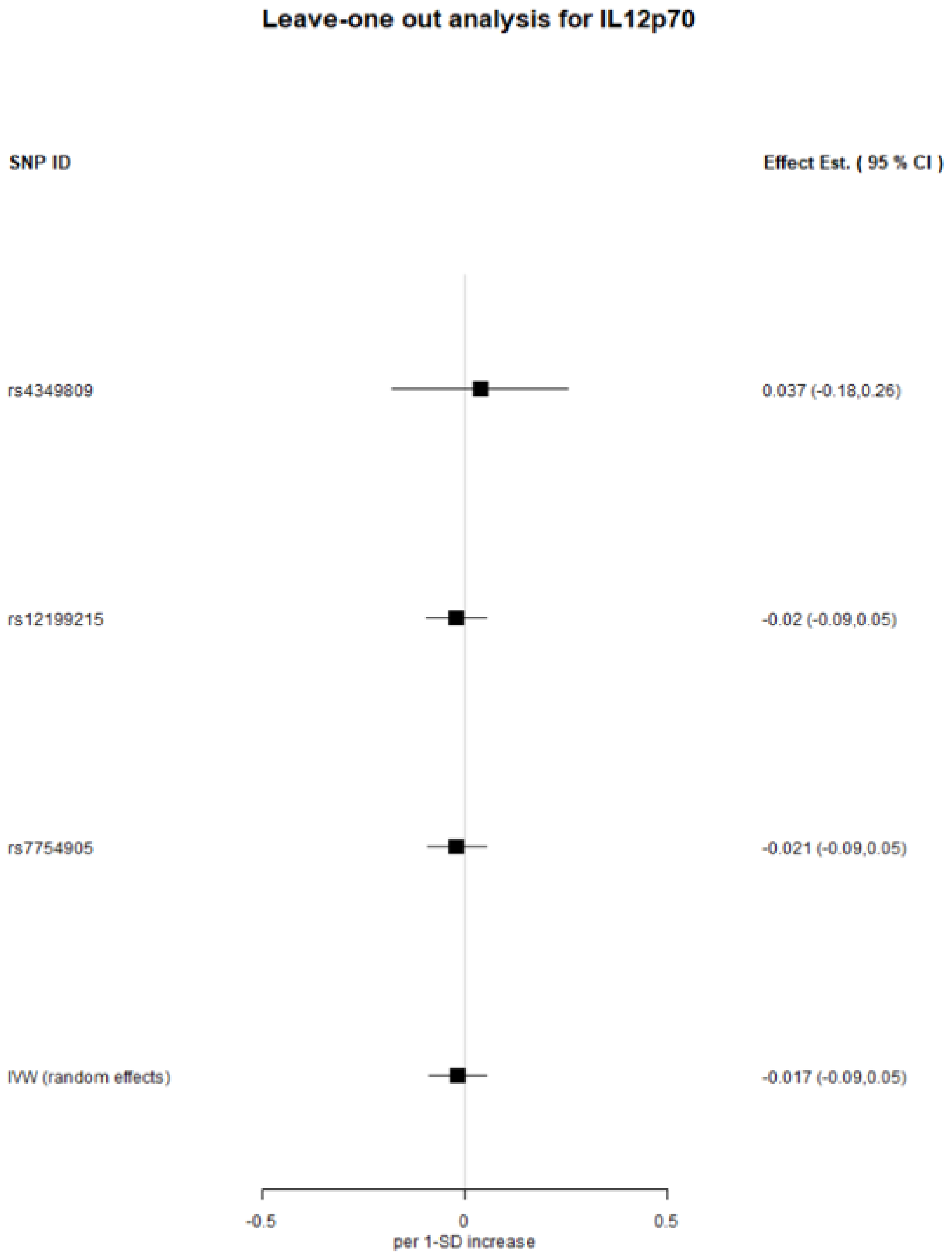


**Figure S8.** Leave-one out analyses of 1-SD increase in IL12p70 on Alzheimer's disease, using IVW. Estimates shown as log odds and 95%CI

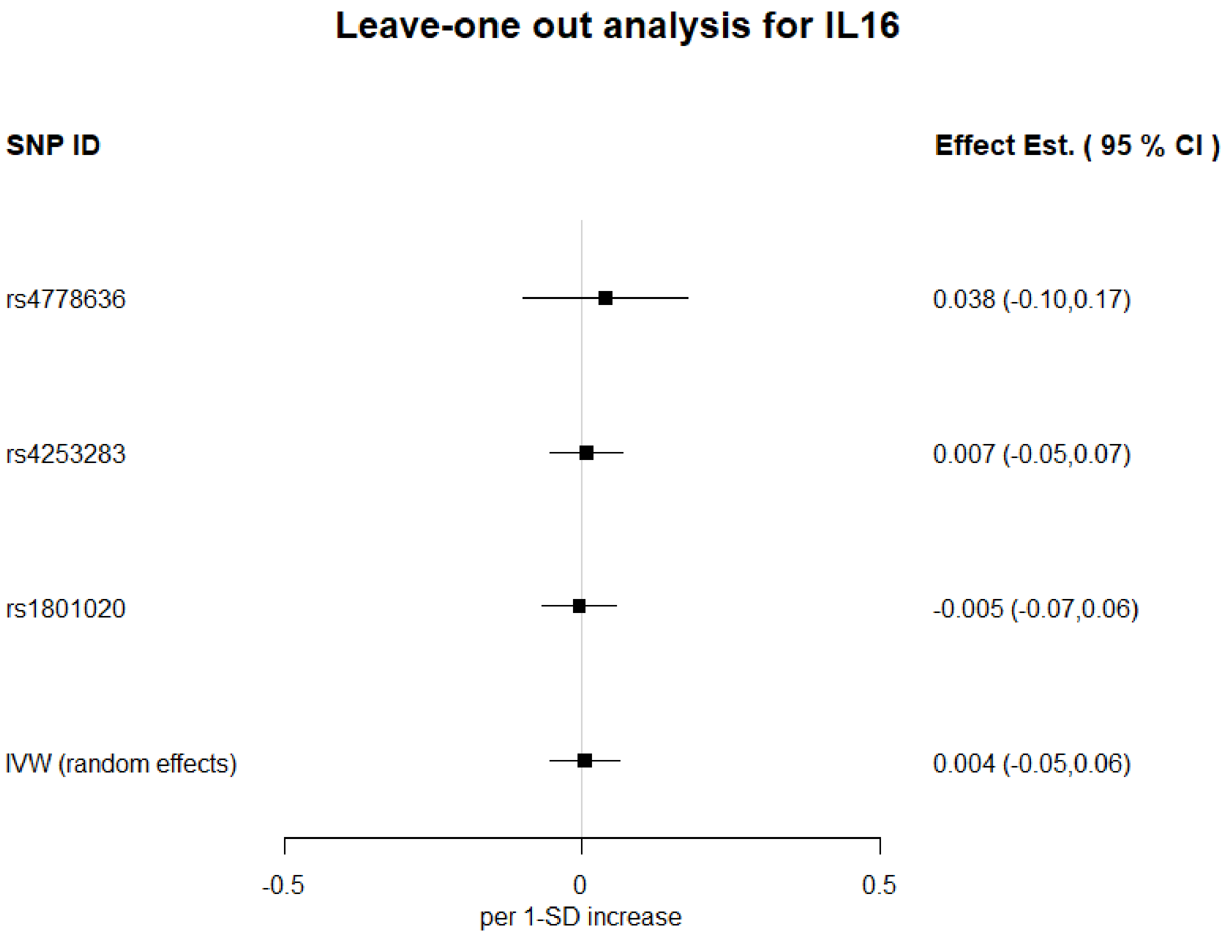


**Figure S9.** Leave-one out analyses of 1-SD increase in IL-16 on Alzheimer's disease, using IVW. Estimates shown as log odds and 95%CI

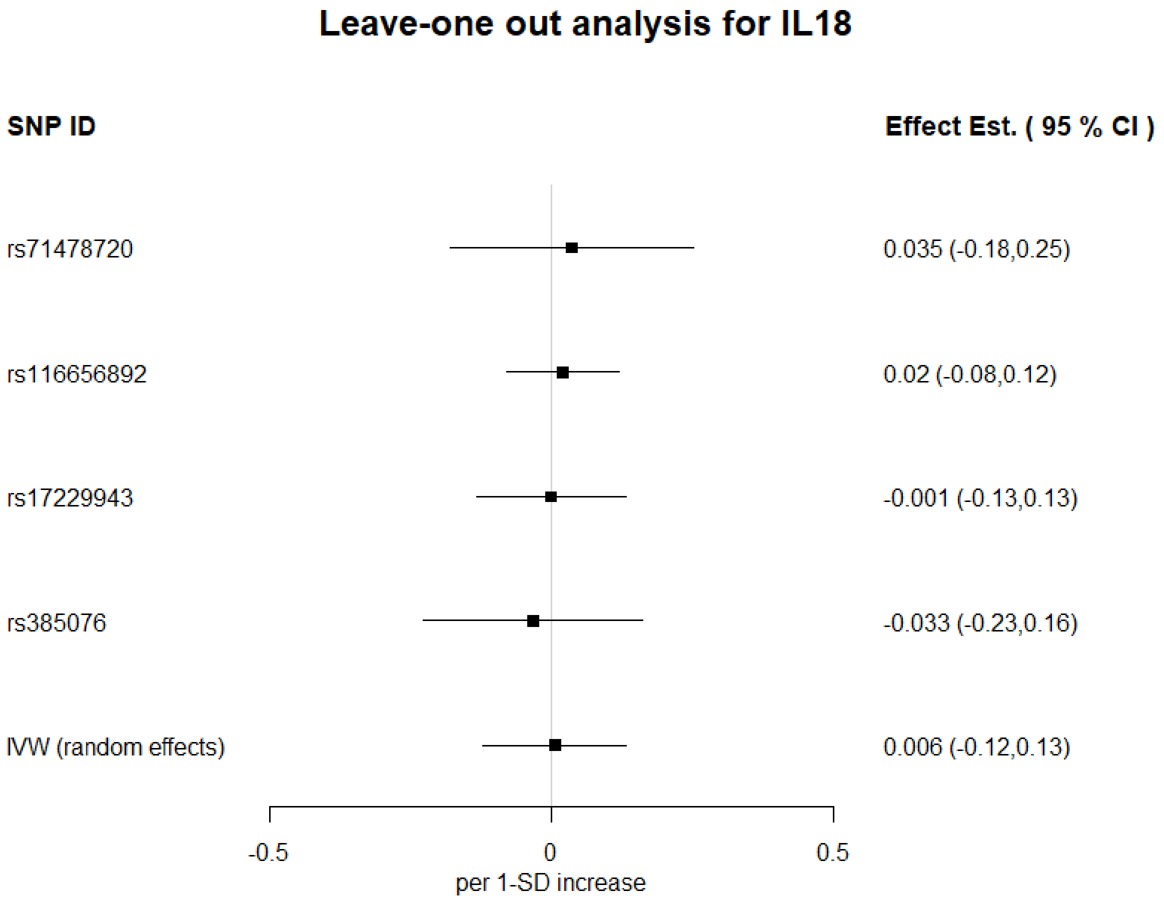


**Figure S10.** Leave-one out analyses of 1-SD increase in IL-18 on Alzheimer's disease, using IVW. Estimates shown as log odds and 95%CI

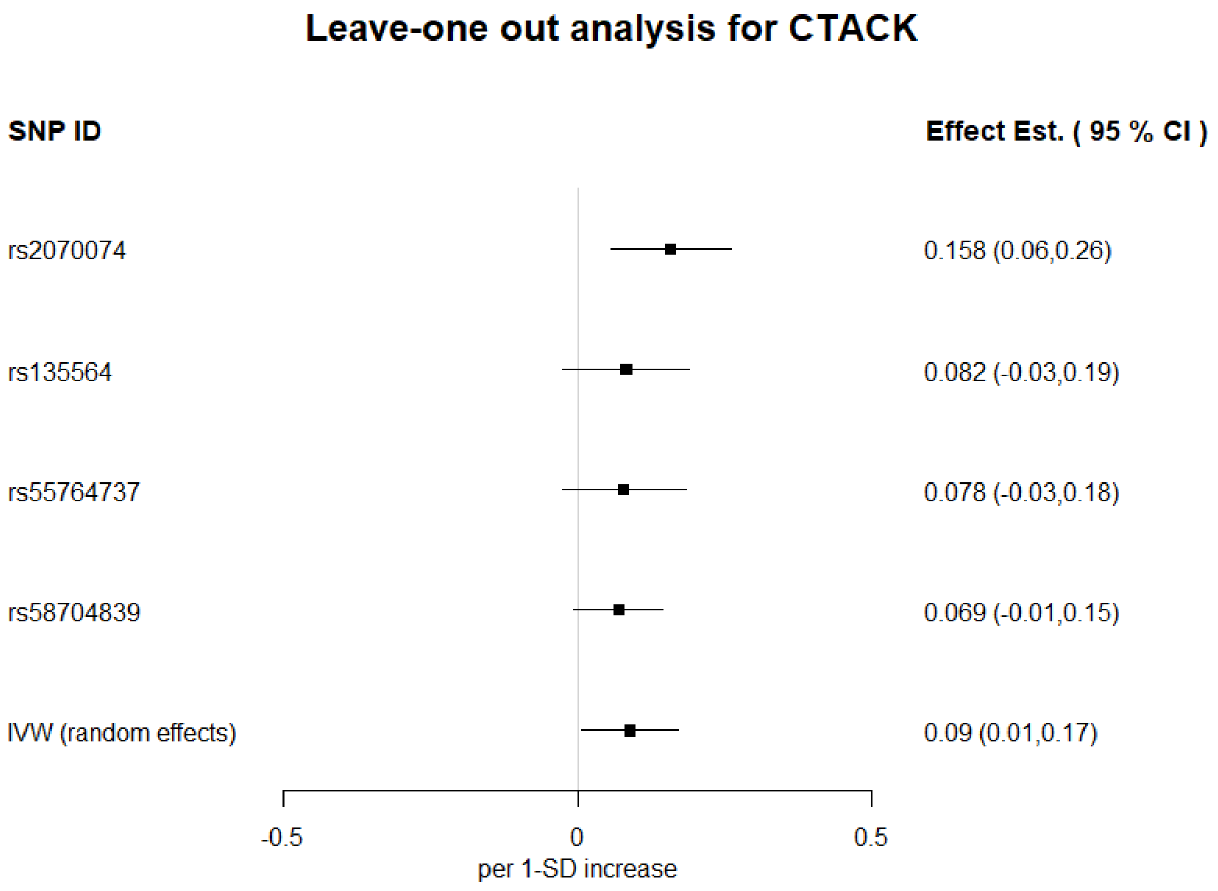


**Figure S11.** Leave-one out analyses of 1-SD increase in CTACK on Alzheimer's disease, using IVW. Estimates shown as log odds and 95%CI

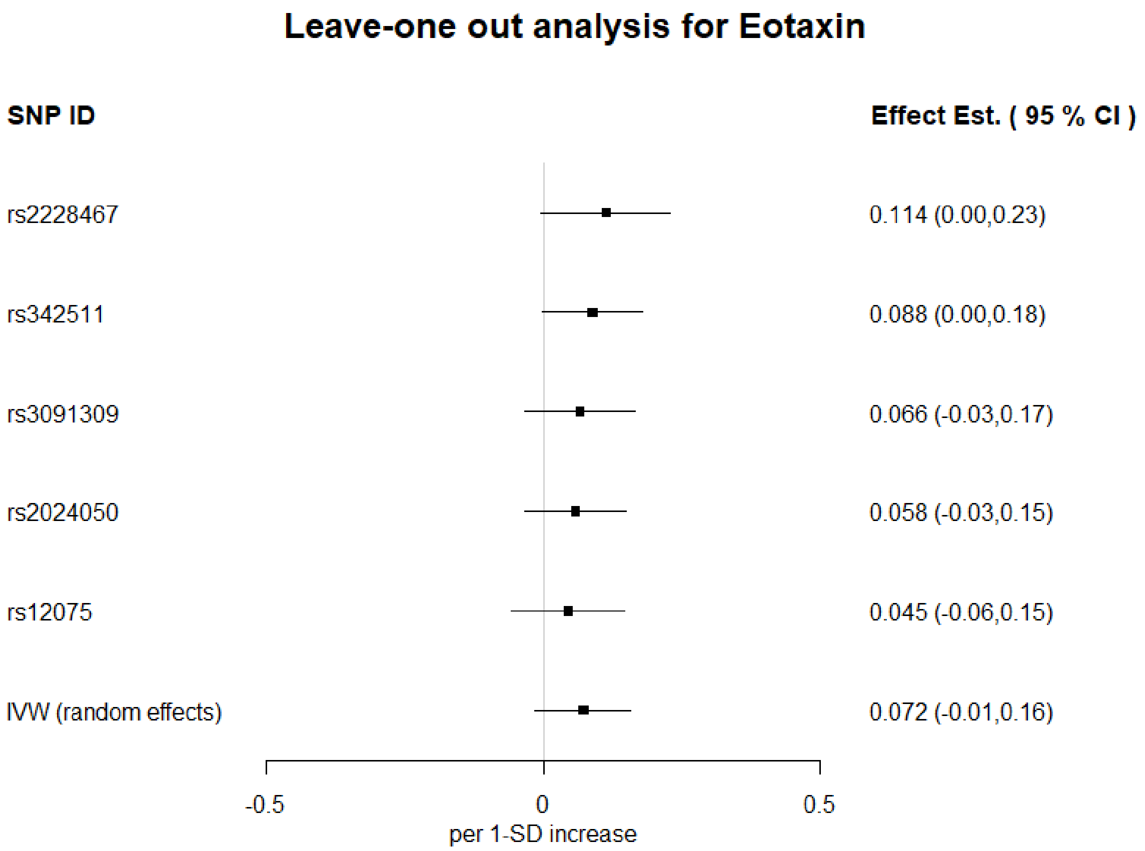


**Figure S12.** Leave-one out analyses of 1-SD increase in Eotaxin on Alzheimer's disease, using IVW. Estimates shown as log odds and 95%CI

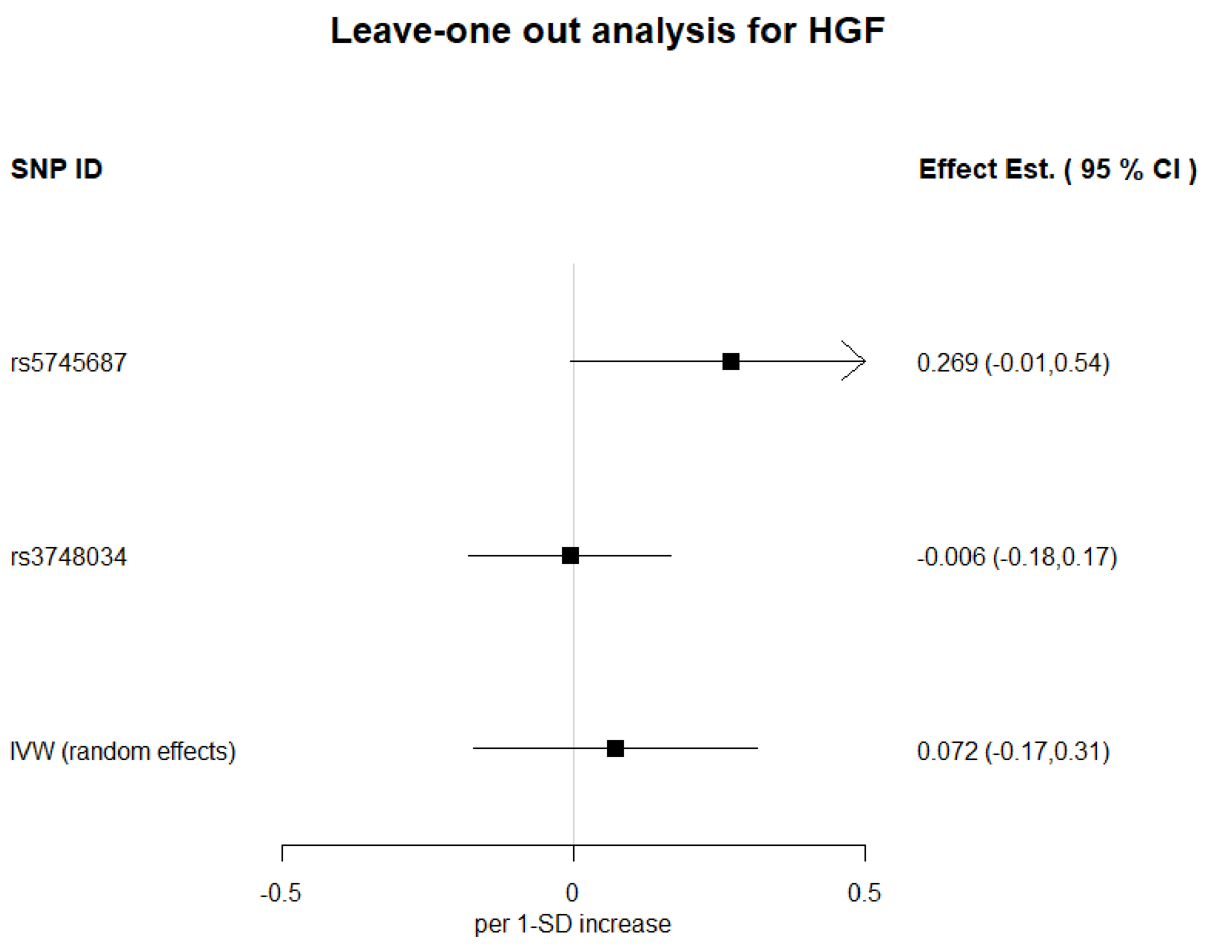


**Figure S13.** Leave-one out analyses of 1-SD increase in HGF on Alzheimer's disease, using IVW. Estimates shown as log odds and 95%CI

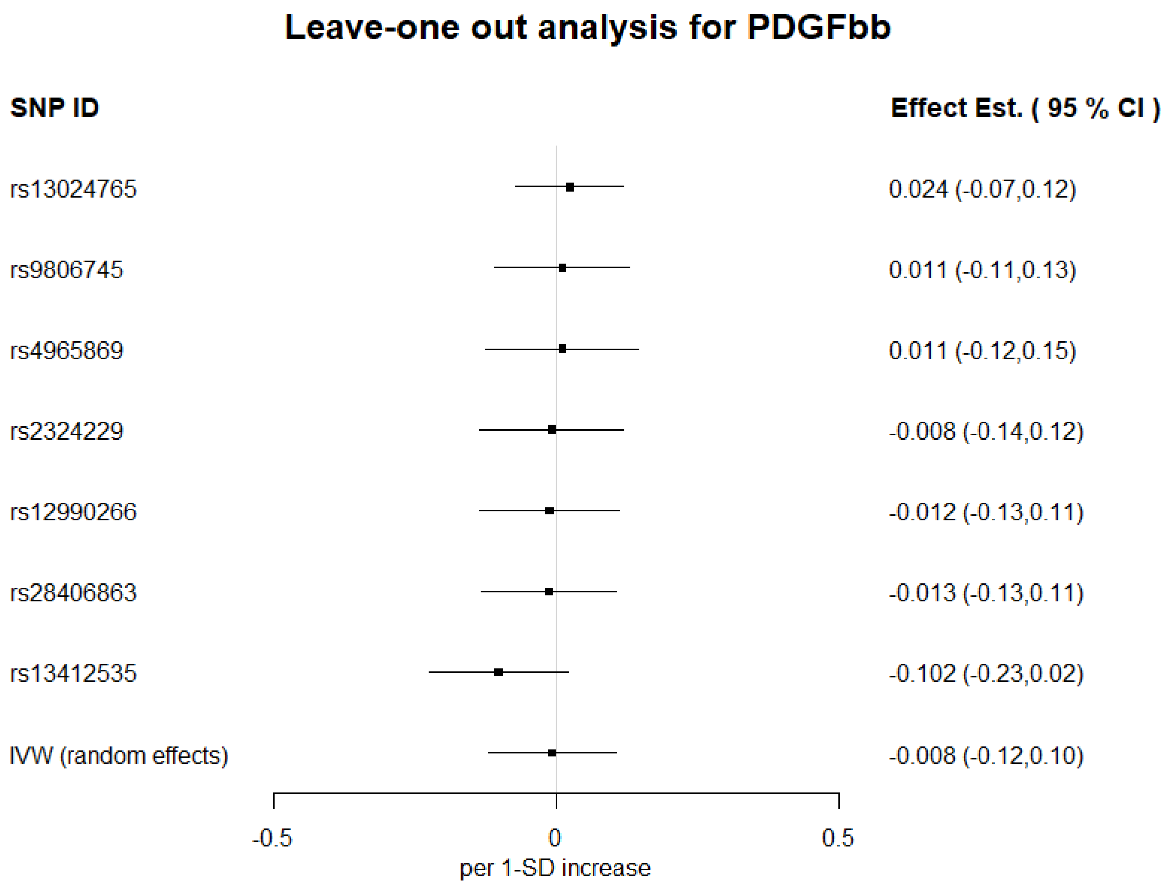


**Figure S14.** Leave-one out analyses of 1-SD increase in PDGFbb on Alzheimer's disease, using IVW. Estimates shown as log odds and 95%CI

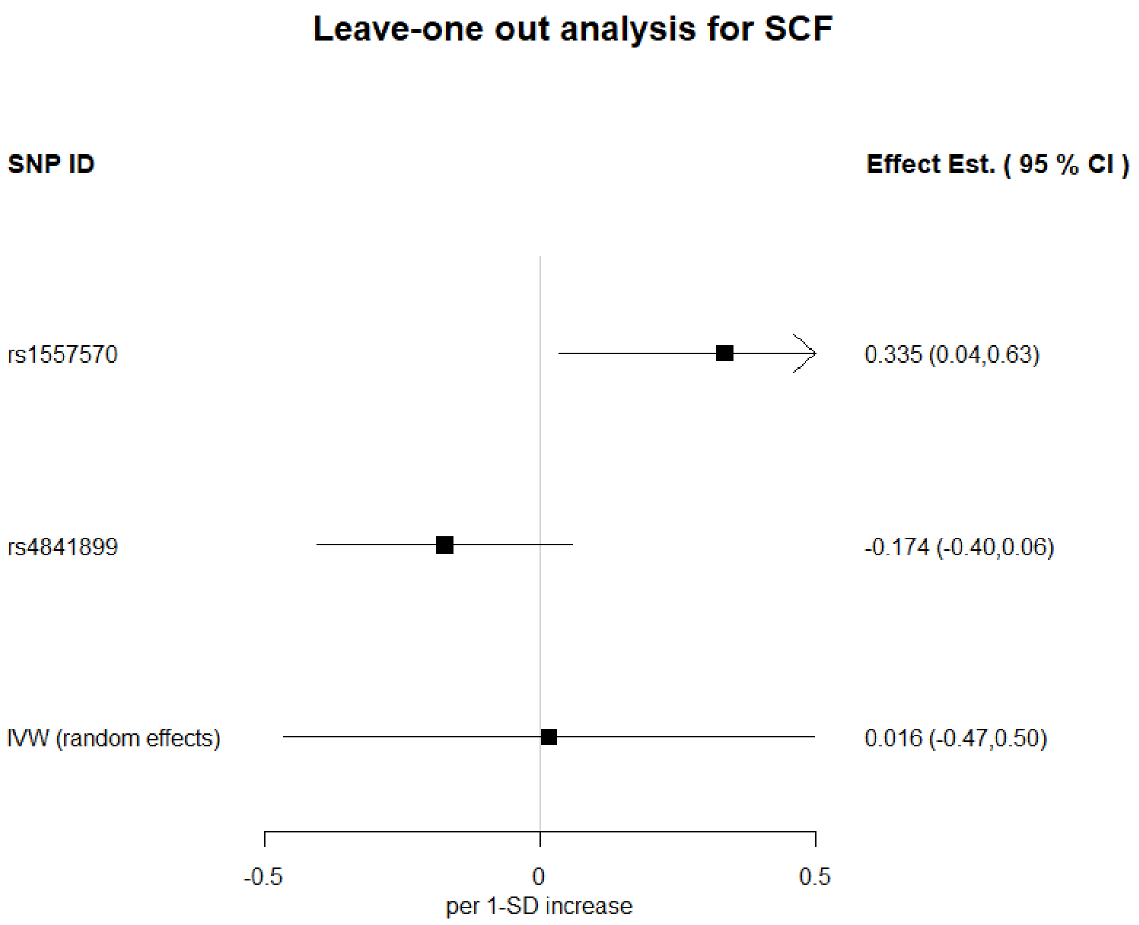


**Figure S15.** Leave-one out analyses of 1-SD increase in SCF on Alzheimer's disease, using IVW. Estimates shown as log odds and 95%CI

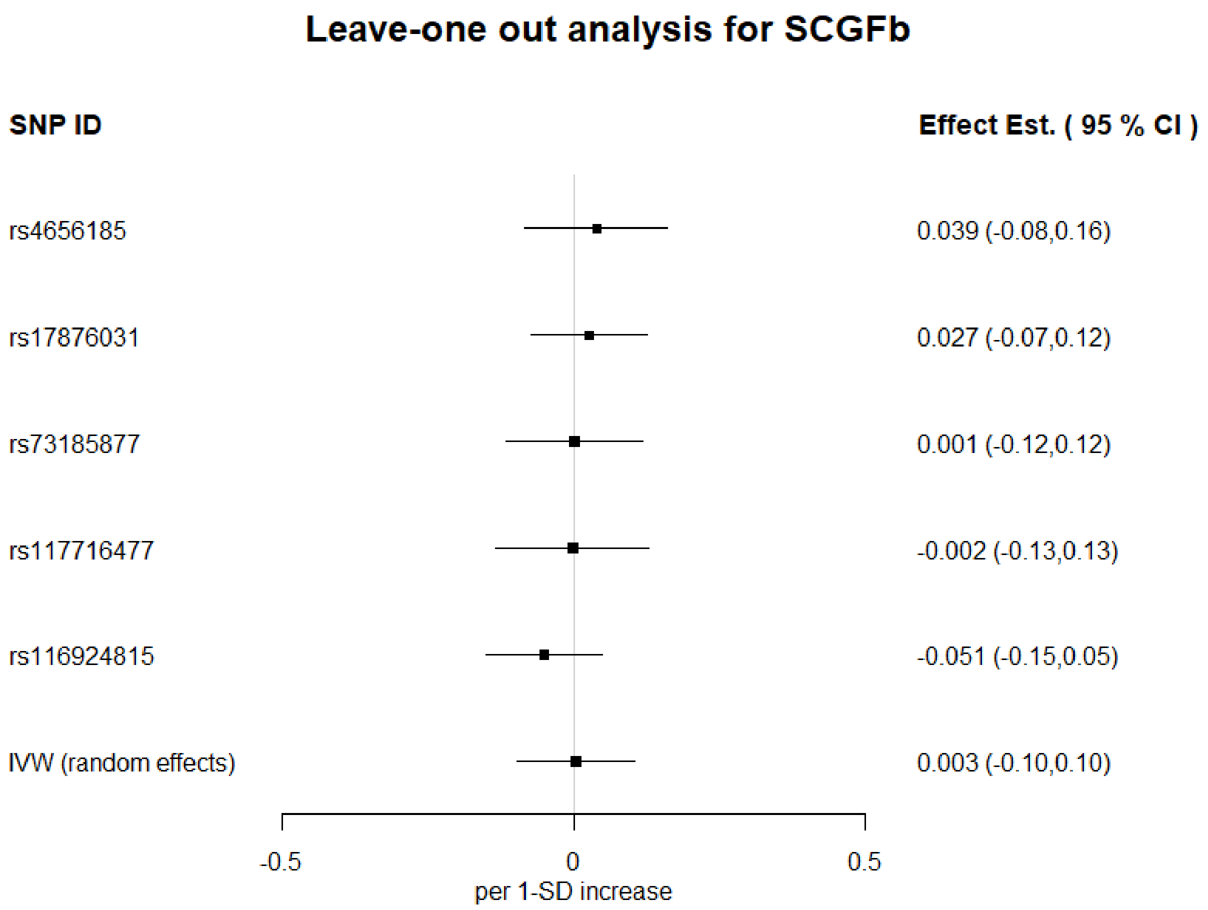


**Figure S16.** Leave-one out analyses of 1-SD increase in SCGFb on Alzheimer's disease, using IVW. Estimates shown as log odds and 95%CI

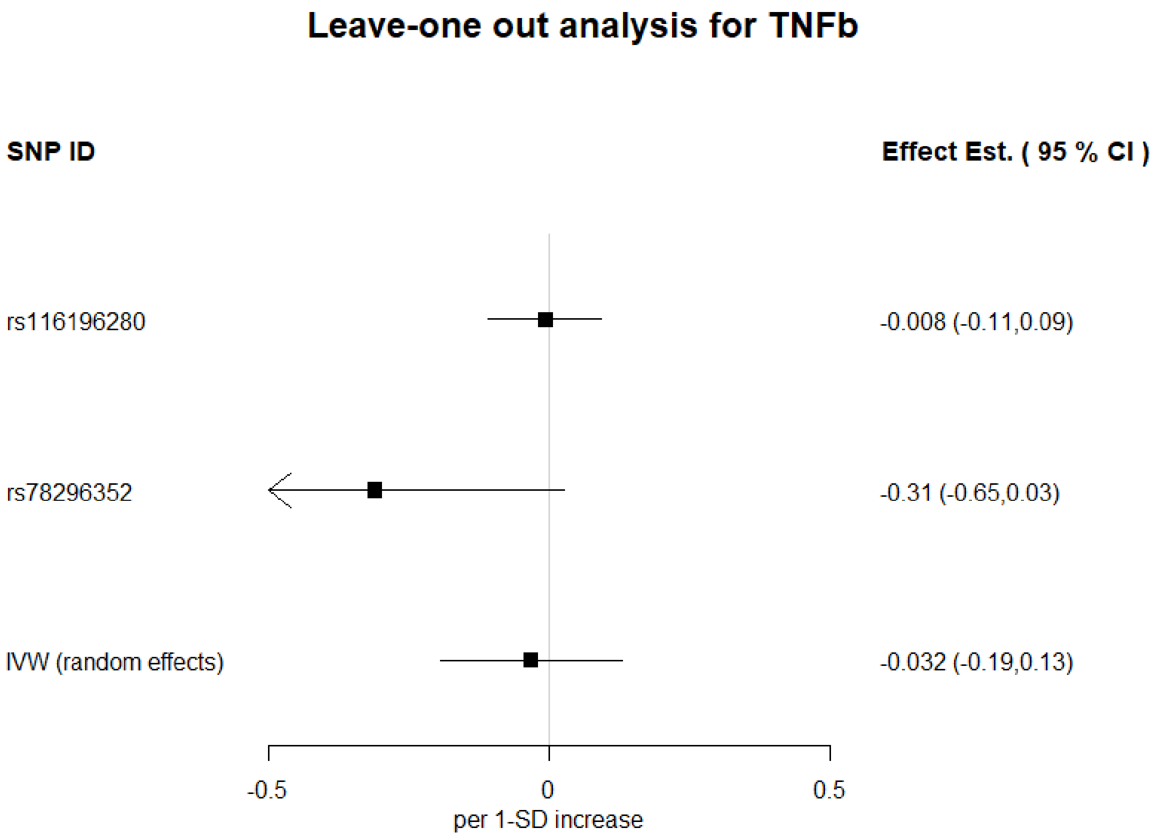


**Figure S17.** Leave-one out analyses of 1-SD increase in TNFb on Alzheimer's disease, using IVW. Estimates shown as log odds and 95%CI

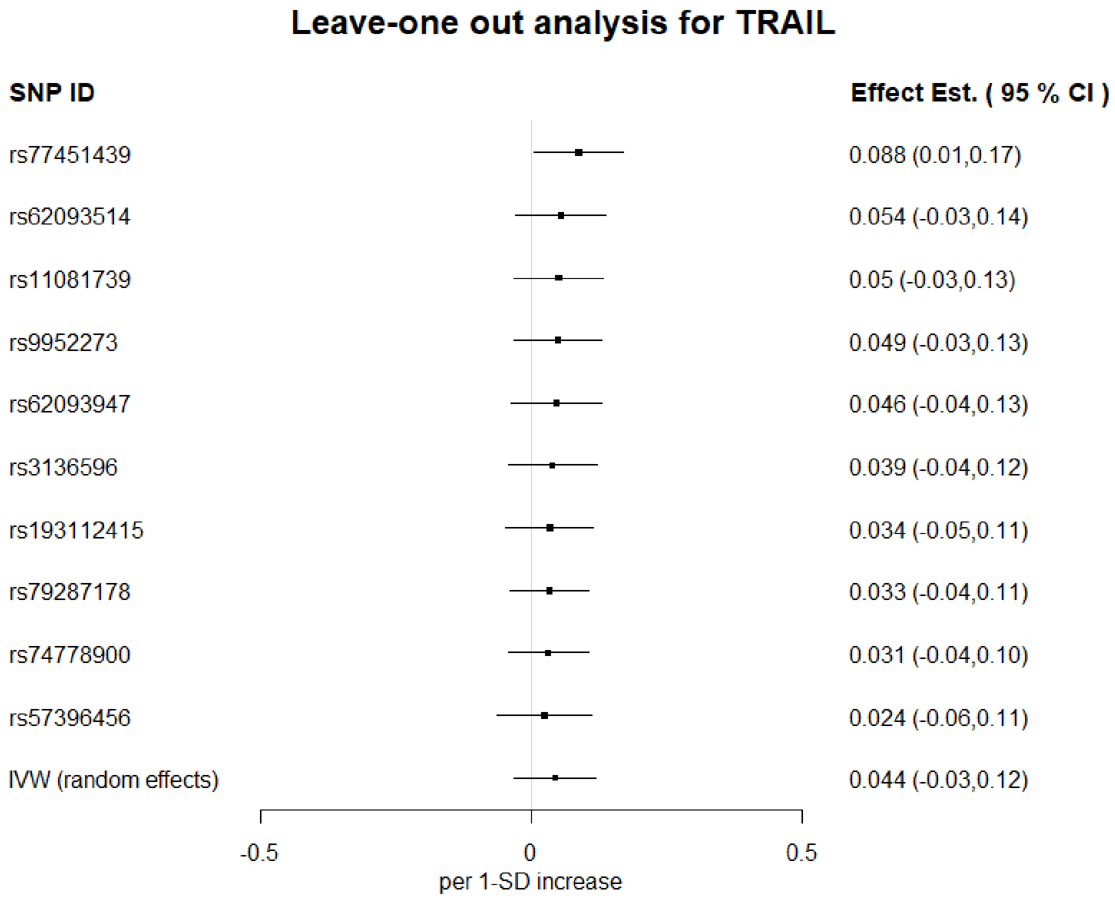


**Figure S18.** Leave-one out analyses of 1-SD increase in TRAIL on Alzheimer's disease, using IVW. Estimates shown as log odds and 95%CI

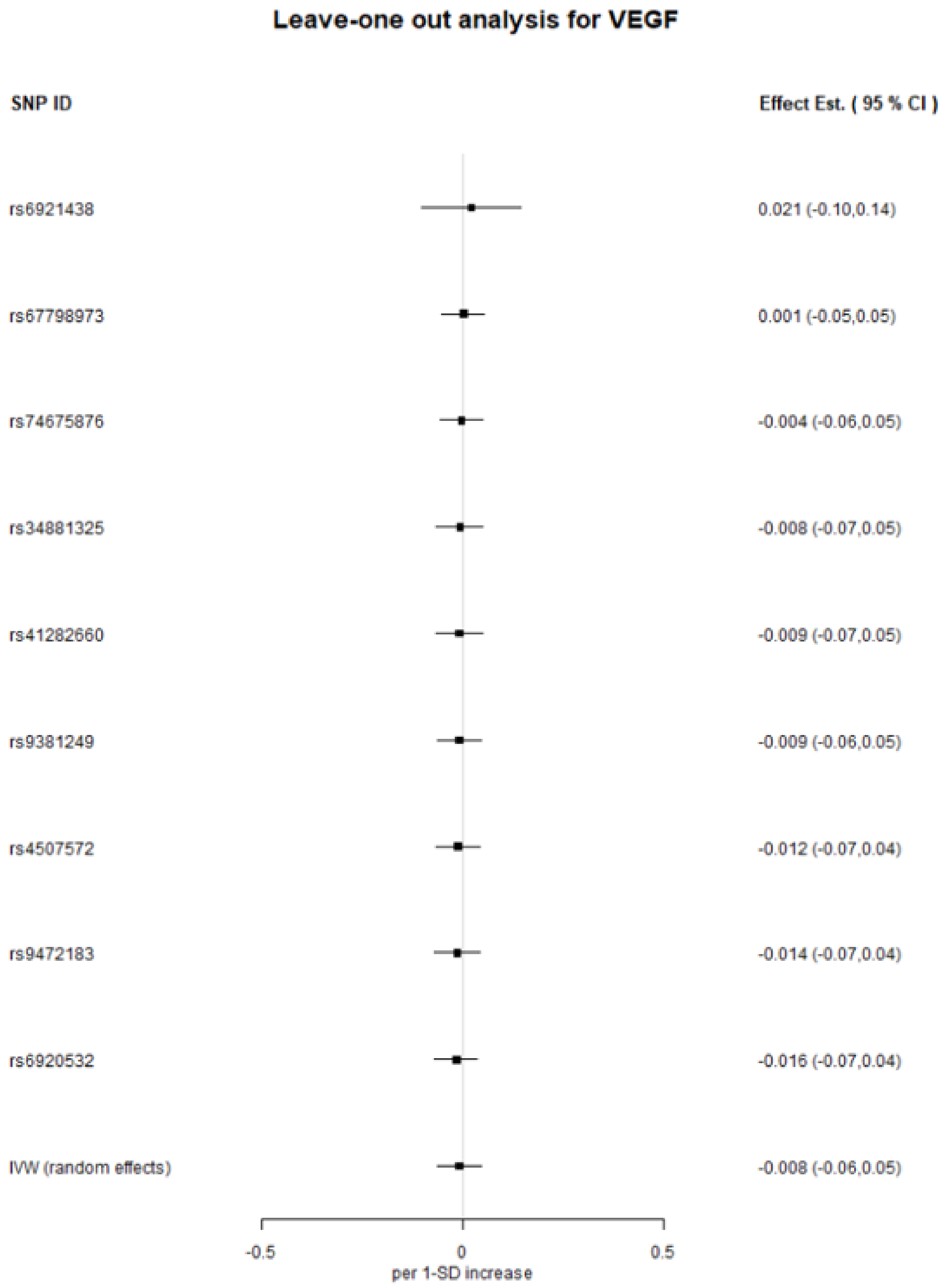


**Figure S19.** Leave-one out analyses of 1-SD increase in VEGF on Alzheimer's disease, using IVW. Estimates shown as log odds and 95%CI

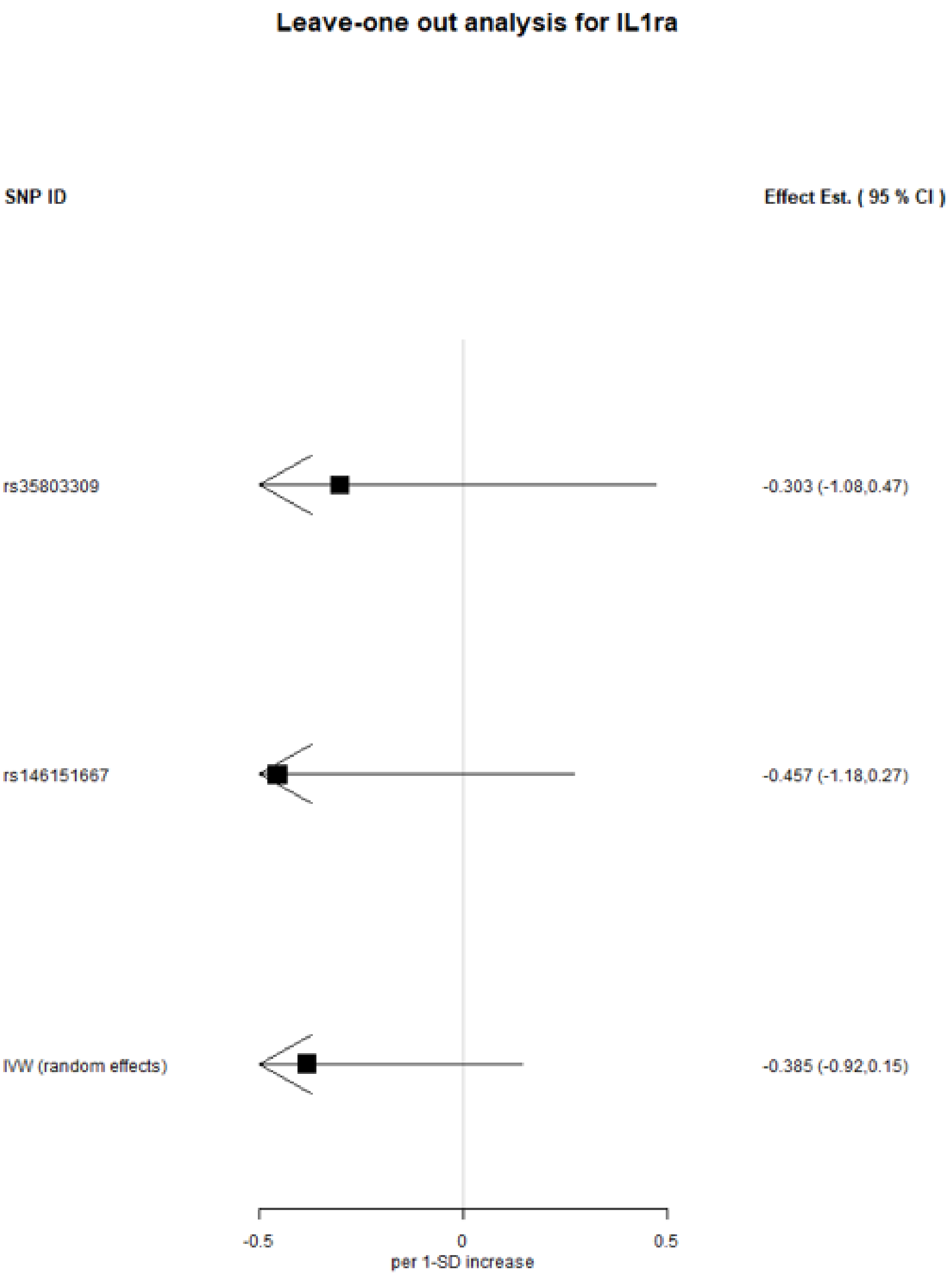


**Figure S20.** Leave-one out analyses of 1-SD increase in IL-1ra on Alzheimer’s disease, using IVW. Estimates shown as log odds and 95%CI

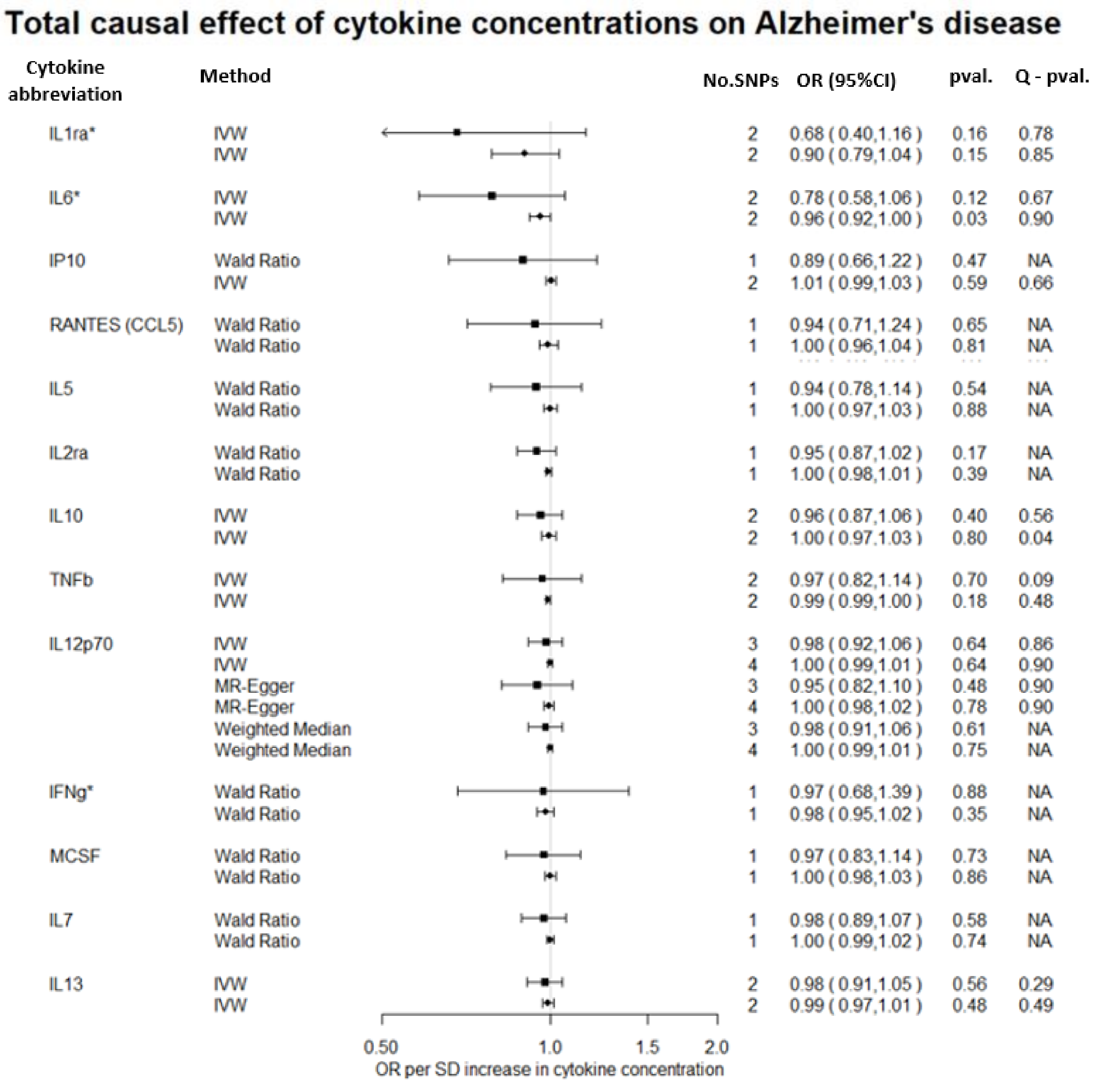


**Figure S21A.** Direct comparison of total causal effects of circulating cytokine concentrations on the risk of Alzheimer's disease, as estimated by Wald Ratio, IVW, MR-Egger and Weighted median estimators, using summary data from Phase 1 (■) and Phase 3 ( ●) of the Alzheimer's GWAS.

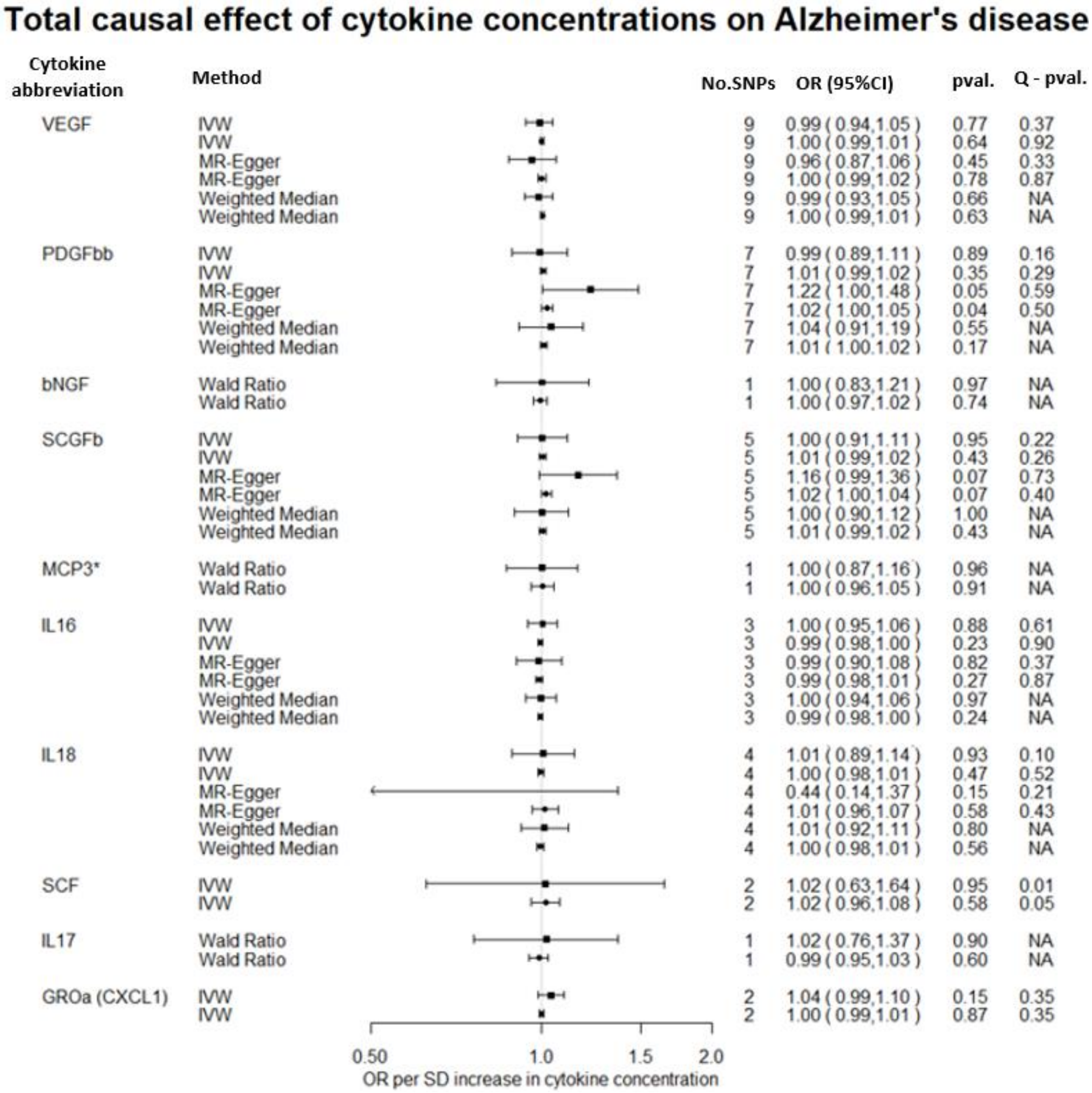


**Figure S21B.** Direct comparison of total causal effects of circulating cytokine concentrations on the risk of Alzheimer’s disease, as estimated by Wald Ratio, IVW, MR-Egger and Weighted median estimators, using summary data from Phase 1 (■) and Phase 3 ( ●) of the Alzheimer’s GWAS.

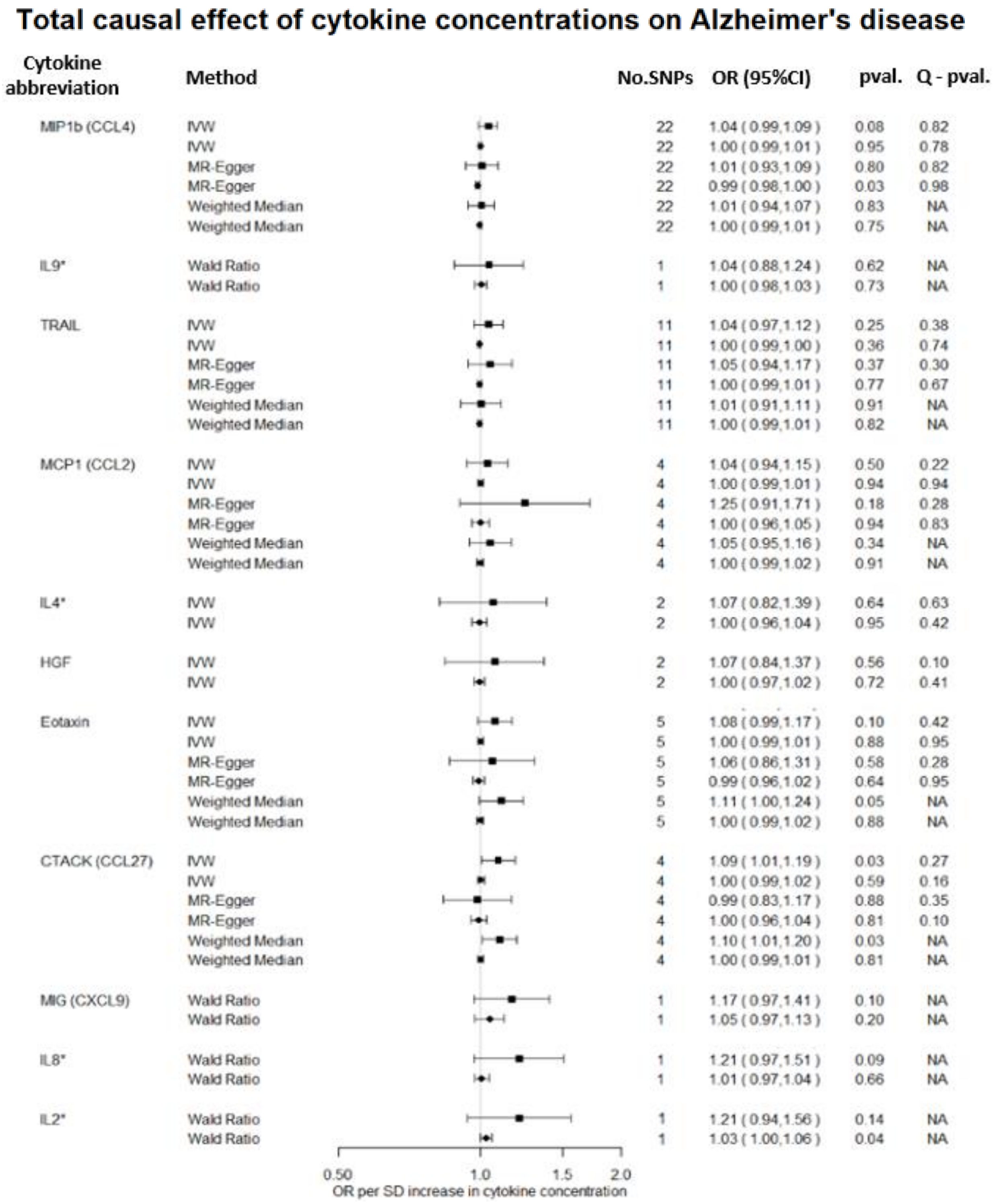


**Figure S21C.** Direct comparison of total causal effects of circulating cytokine concentrations on the risk of Alzheimer's disease, as estimated by Wald Ratio, IVW, MR-Egger and Weighted median estimators, using summary data from Phase 1 (■) and Phase 3 ( ●) of the Alzheimer's GWAS.

### Total causal effect of genetic liability to Alzheimer's disease on cytokine concentrations

| Cytokine abbreviation | Method | No.SNPs | β (95%CI) | pval. | Q - pval. |
|---|---|---|---|---|---|
| IL1ra* | IVW | 26 | -0.03 ( -0.09, 0.03 ) | 0.35 | 0.95 |
| | IVW | 29 | 0.01 ( -0.39, 0.42 ) | 0.95 | 0.94 |
| | MR-Egger | 26 | -0.07 ( -0.18, 0.05 ) | 0.28 | 0.94 |
| | MR-Egger | 29 | -0.21 ( -0.91, 0.48 ) | 0.55 | 0.94 |
| | Weighted Median | 26 | -0.05 ( -0.14, 0.04 ) | 0.25 | NA |
| | Weighted Median | 29 | 0.00 ( -0.58, 0.58 ) | 0.99 | NA |
| IP10 | IVW | 26 | -0.04 ( -0.11, 0.04 ) | 0.32 | 0.06 |
| | IVW | 29 | -0.24 ( -0.67, 0.20 ) | 0.29 | 0.23 |
| | MR-Egger | 26 | -0.12 ( -0.26, 0.02 ) | 0.10 | 0.08 |
| | MR-Egger | 29 | -0.17 ( -0.92, 0.59 ) | 0.67 | 0.19 |
| | Weighted Median | 26 | -0.11 ( -0.20, -0.02 ) | 0.02 | NA |
| | Weighted Median | 29 | -0.02 ( -0.62, 0.57 ) | 0.94 | NA |
| RANTES (CCL5) | IVW | 26 | 0.01 ( -0.06, 0.07 ) | 0.85 | 0.98 |
| | IVW | 29 | -0.09 ( -0.50, 0.33 ) | 0.69 | 0.63 |
| | MR-Egger | 26 | -0.06 ( -0.18, 0.06 ) | 0.33 | 0.99 |
| | MR-Egger | 29 | -0.86 ( -1.57, -0.15 ) | 0.02 | 0.90 |
| | Weighted Median | 26 | -0.01 ( -0.09, 0.08 ) | 0.88 | NA |
| | Weighted Median | 29 | -0.35 ( -0.96, 0.26 ) | 0.26 | NA |
| IL5 | IVW | 26 | 0.00 ( -0.06, 0.06 ) | 0.99 | 0.67 |
| | IVW | 29 | 0.10 ( -0.32, 0.53 ) | 0.63 | 0.52 |
| | MR-Egger | 26 | 0.00 ( -0.12, 0.12 ) | 1.00 | 0.61 |
| | MR-Egger | 29 | 0.02 ( -0.72, 0.75 ) | 0.97 | 0.47 |
| | Weighted Median | 26 | 0.00 ( -0.09, 0.09 ) | 0.96 | NA |
| | Weighted Median | 29 | 0.01 ( -0.61, 0.63 ) | 0.98 | NA |
| IL2ra | IVW | 26 | -0.05 ( -0.11, 0.02 ) | 0.19 | 0.15 |
| | IVW | 29 | 0.06 ( -0.41, 0.54 ) | 0.79 | 0.08 |
| | MR-Egger | 26 | -0.12 ( -0.25, 0.01 ) | 0.08 | 0.17 |
| | MR-Egger | 29 | -0.59 ( -1.35, 0.17 ) | 0.13 | 0.18 |
| | Weighted Median | 26 | -0.04 ( -0.14, 0.05 ) | 0.37 | NA |
| | Weighted Median | 29 | -0.24 ( -0.87, 0.39 ) | 0.45 | NA |
| IL10 | IVW | 26 | -0.01 ( -0.06, 0.05 ) | 0.76 | 0.01 |
| | IVW | 29 | 0.00 ( -0.28, 0.28 ) | 0.99 | 0.42 |
| | MR-Egger | 26 | -0.02 ( -0.13, 0.10 ) | 0.76 | 0.01 |
| | MR-Egger | 29 | 0.15 ( -0.33, 0.63 ) | 0.53 | 0.40 |
| | Weighted Median | 26 | 0.01 ( -0.06, 0.08 ) | 0.82 | NA |
| | Weighted Median | 29 | -0.15 ( -0.55, 0.25 ) | 0.47 | NA |
| IL12p70 | IVW | 26 | 0.00 ( -0.05, 0.05 ) | 0.93 | 0.04 |
| | IVW | 29 | 0.13 ( -0.14, 0.39 ) | 0.36 | 0.63 |
| | MR-Egger | 26 | 0.01 ( -0.09, 0.11 ) | 0.85 | 0.03 |
| | MR-Egger | 29 | 0.30 ( -0.15, 0.75 ) | 0.19 | 0.63 |
| | Weighted Median | 26 | 0.04 ( -0.02, 0.11 ) | 0.19 | NA |
| | Weighted Median | 29 | 0.06 ( -0.32, 0.45 ) | 0.74 | NA |

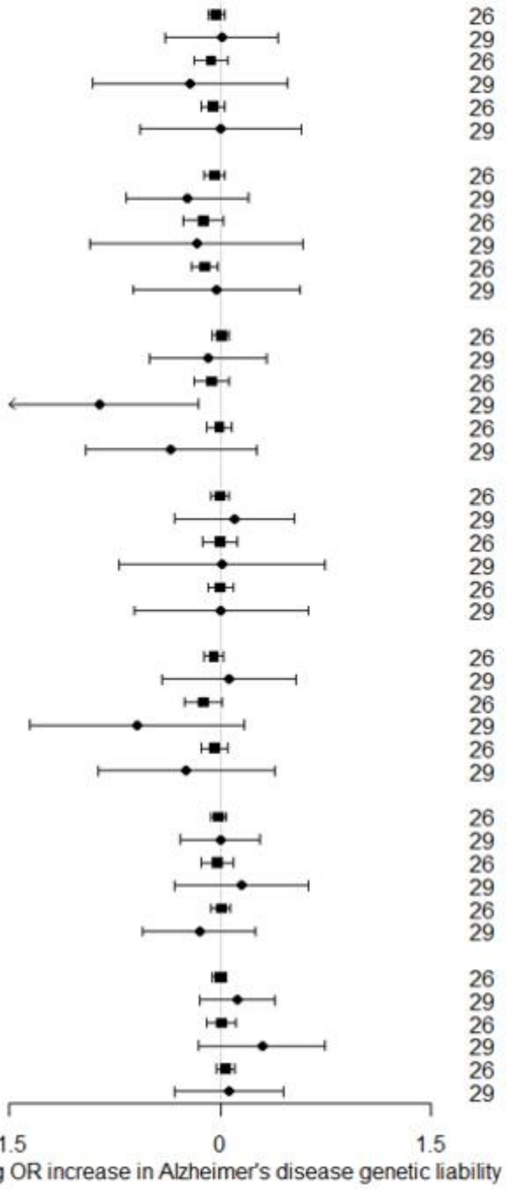


**Figure S22A.** Direct comparison of total causal effect genetic liability to Alzheimer's disease on circulating cytokine concentrations, as estimated by Wald Ratio, IVW, MR-Egger and Weighted median estimators, using summary data from Phase 1 (■) and Phase 3 ( ●) of the Alzheimer's GWAS.

### Total causal effect of genetic liability to Alzheimer's disease on cytokine concentrations

| Cytokine abbreviation | Method | No.SNPs | β (95%CI) | pval. | Q - pval. |
|---|---|---|---|---|---|
| IFNg* | IVW | 26 | -0.01 ( -0.05, 0.03 ) | 0.59 | 0.40 |
| | IVW | 29 | 0.14 ( -0.14, 0.42 ) | 0.32 | 0.59 |
| | MR-Egger | 26 | -0.03 ( -0.12, 0.05 ) | 0.46 | 0.37 |
| | MR-Egger | 29 | 0.22 ( -0.24, 0.69 ) | 0.35 | 0.55 |
| | Weighted Median | 26 | -0.01 ( -0.08, 0.05 ) | 0.71 | NA |
| | Weighted Median | 29 | 0.24 ( -0.18, 0.67 ) | 0.26 | NA |
| MCSF | IVW | 26 | -0.06 ( -0.14, 0.01 ) | 0.10 | 0.62 |
| | IVW | 29 | 0.04 ( -0.50, 0.57 ) | 0.89 | 0.22 |
| | MR-Egger | 26 | 0.00 ( -0.15, 0.14 ) | 0.96 | 0.61 |
| | MR-Egger | 29 | 0.26 ( -0.67, 1.19 ) | 0.58 | 0.20 |
| | Weighted Median | 26 | -0.03 ( -0.14, 0.07 ) | 0.51 | NA |
| | Weighted Median | 29 | 0.22 ( -0.48, 0.92 ) | 0.54 | NA |
| IL7 | IVW | 26 | 0.01 ( -0.06, 0.07 ) | 0.84 | 0.26 |
| | IVW | 29 | 0.28 ( -0.15, 0.71 ) | 0.20 | 0.41 |
| | MR-Egger | 26 | 0.05 ( -0.09, 0.18 ) | 0.49 | 0.24 |
| | MR-Egger | 29 | 0.33 ( -0.42, 1.09 ) | 0.38 | 0.36 |
| | Weighted Median | 26 | 0.04 ( -0.05, 0.14 ) | 0.36 | NA |
| | Weighted Median | 29 | 0.28 ( -0.33, 0.90 ) | 0.36 | NA |
| IL13 | IVW | 26 | -0.01 ( -0.07, 0.05 ) | 0.78 | 0.65 |
| | IVW | 29 | -0.04 ( -0.45, 0.37 ) | 0.85 | 0.96 |
| | MR-Egger | 26 | 0.01 ( -0.11, 0.13 ) | 0.91 | 0.60 |
| | MR-Egger | 29 | 0.00 ( -0.70, 0.70 ) | 1.00 | 0.94 |
| | Weighted Median | 26 | 0.03 ( -0.07, 0.12 ) | 0.57 | NA |
| | Weighted Median | 29 | -0.06 ( -0.65, 0.54 ) | 0.85 | NA |
| VEGF | IVW | 26 | -0.02 ( -0.08, 0.05 ) | 0.61 | 0.00 |
| | IVW | 29 | -0.07 ( -0.39, 0.25 ) | 0.66 | 0.20 |
| | MR-Egger | 26 | 0.02 ( -0.10, 0.15 ) | 0.71 | 0.00 |
| | MR-Egger | 29 | 0.47 ( -0.02, 0.96 ) | 0.06 | 0.47 |
| | Weighted Median | 26 | 0.03 ( -0.04, 0.10 ) | 0.41 | NA |
| | Weighted Median | 29 | 0.13 ( -0.32, 0.57 ) | 0.58 | NA |
| PDGFbb | IVW | 26 | -0.01 ( -0.05, 0.04 ) | 0.80 | 0.43 |
| | IVW | 29 | -0.02 ( -0.29, 0.25 ) | 0.88 | 0.75 |
| | MR-Egger | 26 | -0.01 ( -0.09, 0.07 ) | 0.83 | 0.38 |
| | MR-Egger | 29 | -0.02 ( -0.47, 0.43 ) | 0.94 | 0.71 |
| | Weighted Median | 26 | 0.03 ( -0.03, 0.08 ) | 0.37 | NA |
| | Weighted Median | 29 | -0.11 ( -0.49, 0.27 ) | 0.56 | NA |
| bNGF | IVW | 26 | -0.01 ( -0.08, 0.06 ) | 0.72 | 0.15 |
| | IVW | 29 | 0.15 ( -0.30, 0.60 ) | 0.52 | 0.20 |
| | MR-Egger | 26 | 0.00 ( -0.14, 0.14 ) | 1.00 | 0.12 |
| | MR-Egger | 29 | 0.02 ( -0.76, 0.81 ) | 0.95 | 0.17 |
| | Weighted Median | 26 | -0.03 ( -0.13, 0.06 ) | 0.50 | NA |
| | Weighted Median | 29 | -0.18 ( -0.82, 0.45 ) | 0.57 | NA |

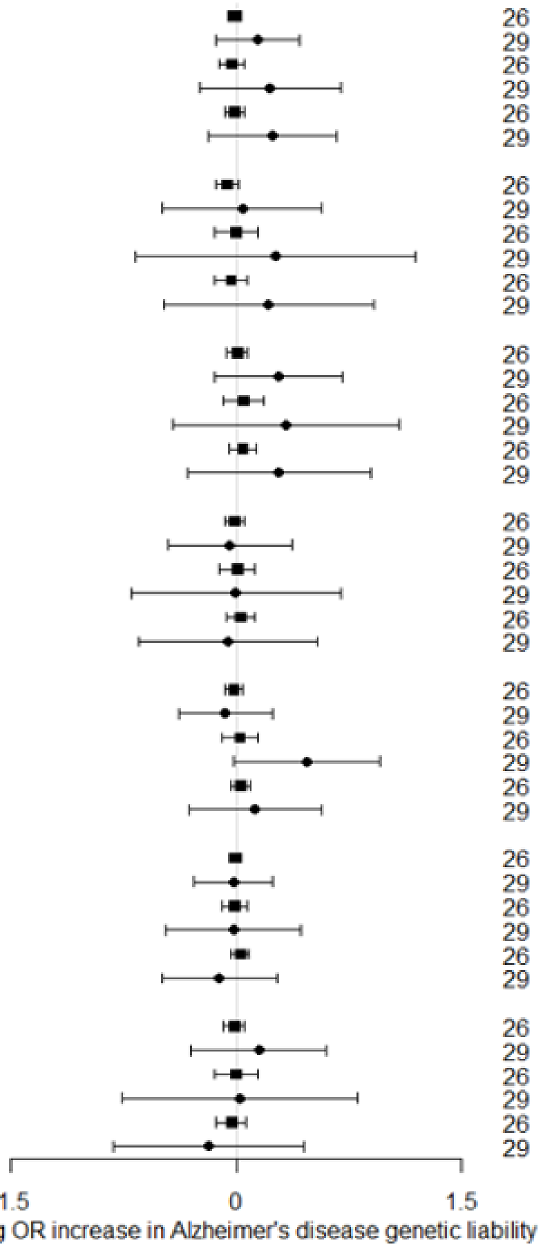


**Figure S22B.** Direct comparison of total causal effect genetic liability to Alzheimer's disease on circulating cytokine concentrations, as estimated by Wald Ratio, IVW, MR-Egger and Weighted median estimators, using summary data from Phase 1 (■) and Phase 3 ( ●) of the Alzheimer's GWAS.

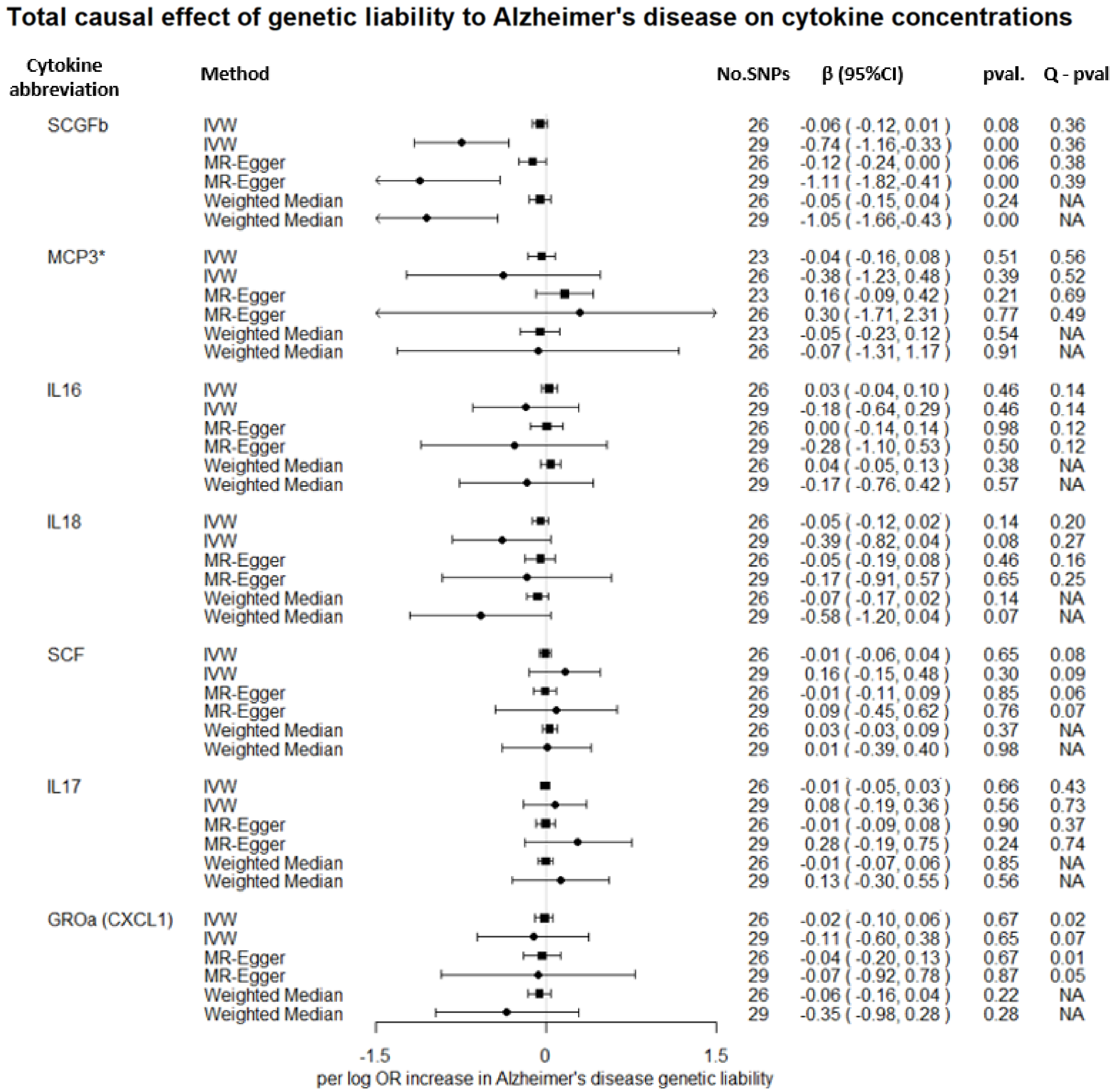


**Figure S22C.** Direct comparison of total causal effect genetic liability to Alzheimer's disease on circulating cytokine concentrations, as estimated by Wald Ratio, IVW, MR-Egger and Weighted median estimators, using summary data from Phase 1 (■) and Phase 3 ( ●) of the Alzheimer's GWAS.

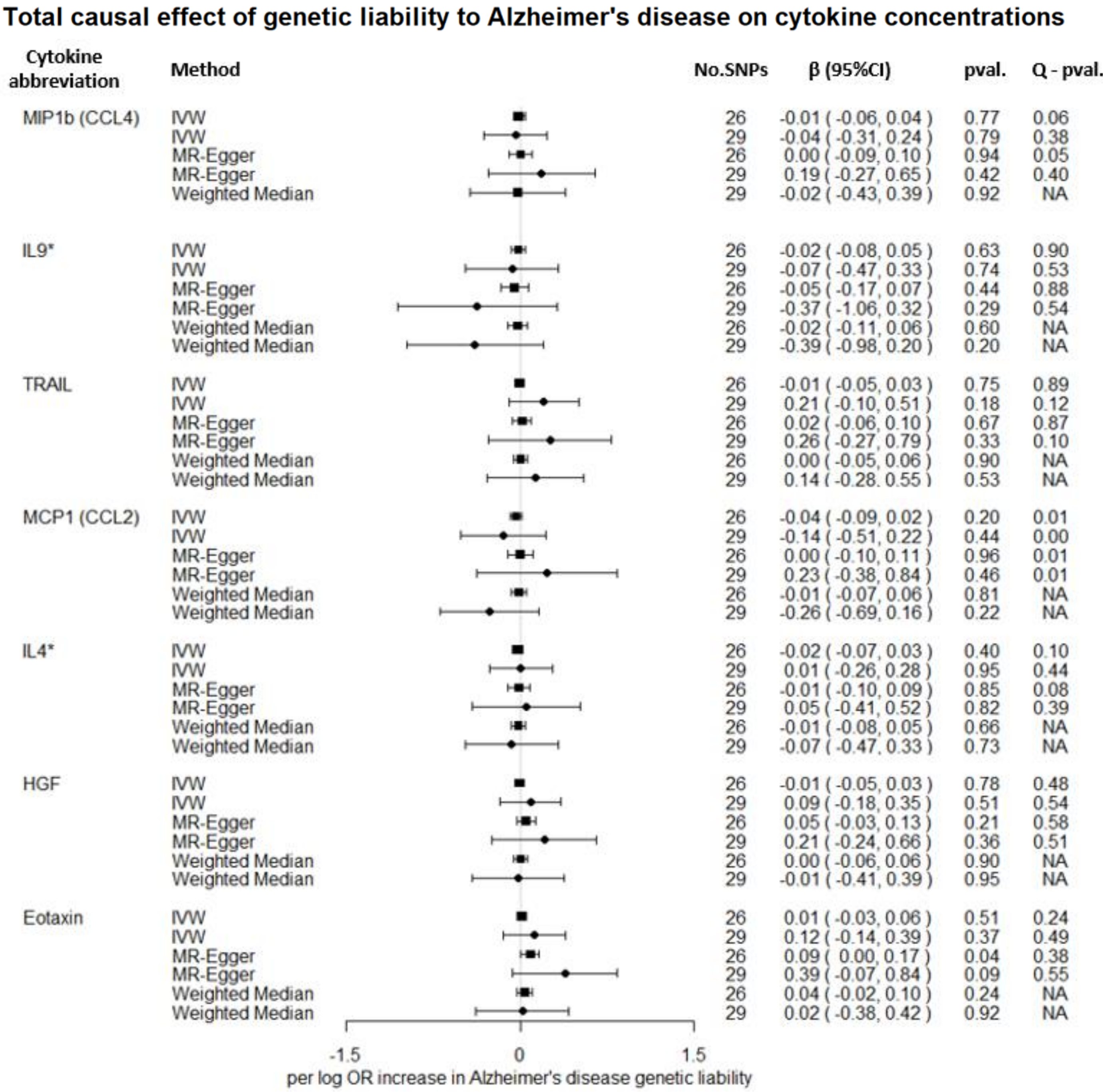


**Figure S22D.** Direct comparison of total causal effect genetic liability to Alzheimer's disease on circulating cytokine concentrations, as estimated by Wald Ratio, IVW, MR-Egger and Weighted median estimators, using summary data from Phase 1 (■) and Phase 3 ( ●) of the Alzheimer's GWAS.

**Total causal effect of genetic liability to Alzheimer's disease on cytokine concentrations**

| Cytokine abbreviation | Method | No.SNPs | β (95%CI) | pval. | Q - pval. |
|---|---|---|---|---|---|
| CTACK (CCL27) | IVW | 26 | -0.04 ( -0.11, 0.02 ) | 0.20 | 0.19 |
| | IVW | 29 | -0.24 ( -0.65, 0.16 ) | 0.24 | 0.61 |
| | MR-Egger | 26 | 0.00 ( -0.14, 0.13 ) | 0.95 | 0.17 |
| | MR-Egger | 29 | 0.04 ( -0.65, 0.74 ) | 0.90 | 0.61 |
| | Weighted Median | 26 | -0.03 ( -0.13, 0.06 ) | 0.47 | NA |
| | Weighted Median | 29 | -0.20 ( -0.79, 0.38 ) | 0.50 | NA |
| MIG (CXCL9) | IVW | 26 | -0.03 ( -0.09, 0.03 ) | 0.31 | 0.58 |
| | IVW | 29 | -0.12 ( -0.53, 0.28 ) | 0.55 | 0.43 |
| | MR-Egger | 26 | -0.04 ( -0.16, 0.07 ) | 0.46 | 0.53 |
| | MR-Egger | 29 | -0.33 ( -1.02, 0.37 ) | 0.36 | 0.40 |
| | Weighted Median | 26 | -0.07 ( -0.16, 0.02 ) | 0.10 | NA |
| | Weighted Median | 29 | -0.18 ( -0.79, 0.43 ) | 0.57 | NA |
| IL8* | IVW | 26 | -0.05 ( -0.12, 0.02 ) | 0.14 | 0.20 |
| | IVW | 29 | 0.18 ( -0.24, 0.59 ) | 0.40 | 0.85 |
| | MR-Egger | 26 | -0.05 ( -0.19, 0.08 ) | 0.46 | 0.16 |
| | MR-Egger | 29 | 0.24 ( -0.46, 0.95 ) | 0.50 | 0.82 |
| | Weighted Median | 26 | -0.07 ( -0.17, 0.02 ) | 0.14 | NA |
| | Weighted Median | 29 | 0.02 ( -0.58, 0.62 ) | 0.95 | NA |
| IL2* | IVW | 26 | 0.04 ( -0.02, 0.10 ) | 0.20 | 0.90 |
| | IVW | 29 | 0.33 ( -0.08, 0.75 ) | 0.11 | 0.96 |
| | MR-Egger | 26 | 0.04 ( -0.08, 0.16 ) | 0.54 | 0.87 |
| | MR-Egger | 29 | 0.16 ( -0.56, 0.88 ) | 0.66 | 0.96 |
| | Weighted Median | 26 | 0.05 ( -0.04, 0.14 ) | 0.27 | NA |
| | Weighted Median | 29 | 0.22 ( -0.37, 0.81 ) | 0.46 | NA |

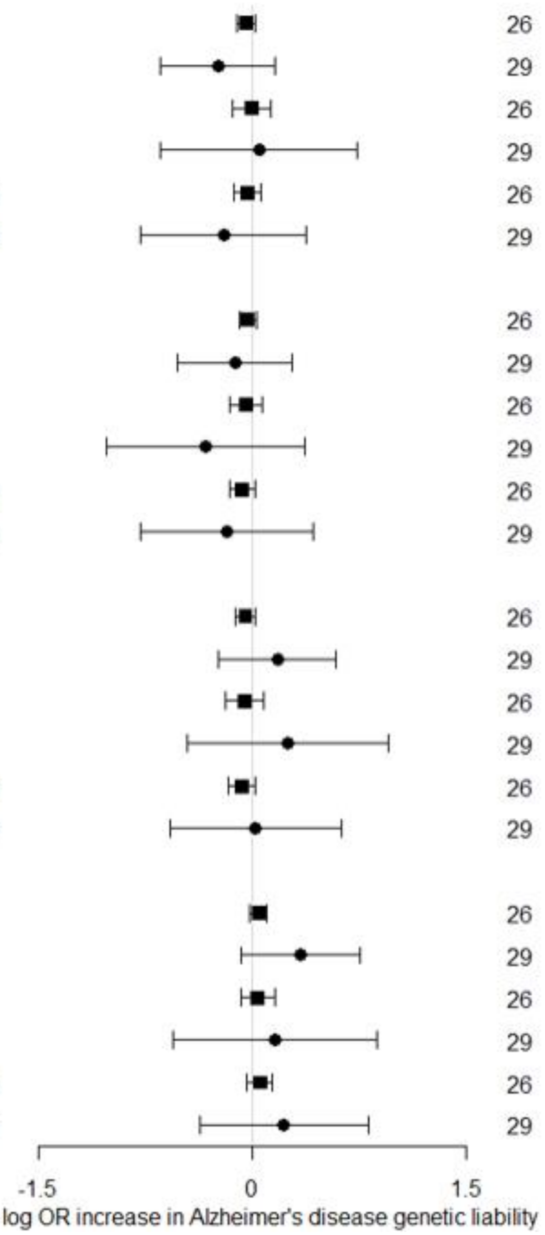


**Figure S22E.** Direct comparison of total causal effect genetic liability to Alzheimer's disease on circulating cytokine concentrations, as estimated by Wald Ratio, IVW, MR-Egger and Weighted median estimators, using summary data from Phase 1 (■) and Phase 3 ( ●) of the Alzheimer's GWAS.
